# The genetic landscape of heterogeneity in human functional brain connectivity

**DOI:** 10.1101/2025.04.08.25325384

**Authors:** Bernardo de APC Maciel, Marijn Schipper, Cato Romero, Christiaan de Leeuw, Koen Helwegen, Danielle Posthuma, Jeanne E. Savage, Martijn P. van den Heuvel

## Abstract

Understanding the genetic underpinnings of functional brain connectivity is essential to understand its role in brain health and disease. A mass-univariate approach was adopted to investigate the genetic architecture of functional brain circuitry (UKB, n_total_ = 28,159 subjects), showing common genetic variants in 33% of all 3,321 interregional functional pathways. Seventy-two locus-connection associations with widespread (pleiotropic) effects throughout the brain were found and mapped to five protein-coding genes: *PAX8, EphA3, SLC39A12, THBS1* and *APOE*. Functional annotation of these genes revealed converged in biological processes related to neurodevelopment, alongside with significant overlap with phenotypes in cardiovascular and cognitive phenotypes domains (enrichment minimum p = 3.0·10^-6^ and p = 1.6·10^-5^, respectively). Our findings show a genetic component of interindividual differences in functional brain connectivity that is shared with traits related to cognitive function and overall health.

## Introduction

Synchronization in neural activity between regions is observed throughout the human brain.^1^ These macroscale dynamic activity patterns form distinct functional networks of synchronous activity across multiple neural systems. These functional connections are important for healthy brain functioning, cognition and behaviour.^2–4^ Functional connectivity (FC) also carries a strong genetic component. Twin and family studies show high heritability of functional networks^5–7^ and recent large-scale Genome-Wide Association Studies (GWAS) have identified relevant genetic loci underlying this high heritability.^8–12^

Several genes have already been identified to play an important role in macroscale brain properties. Activity of the default mode network – a central brain network involved in cognition, self-awareness and theory of mind – is associated with the *APOE* gene, a well-known genetic risk factor for mortality, brain atrophy and Alzheimer’s disease.^8,13,14^ Lifetime trajectories of lifetime brain atrophy are associated with *GPR139*, *DACH1* and *APOE* genes, all of which partake in metabolic processes relevant both in early brain development and neurodegeneration.^13^ *TOMM40*, *APOE* and *APOC1* are associated with FC of brain networks, showing genetic factors with widespread effects throughout the brain.^10^ Moreover, genetic associations are often shared across neuropsychiatric^15–17^ and MRI-derived (magnetic resonance imaging)^17–19^ traits, pointing towards common biological processes driving changes in these phenotypes and suggesting that genetic differences might underlie heterogeneity, i.e. interindividual differences, in FC. Understanding the extent to which specific genetic variants affect different functional brain circuitry can provide mechanistic insight into brain functioning in health and disease.

In this study, we characterise the global genetic architecture of human functional brain connectivity by performing mass-univariate GWAS of all 3,321 pairwise connections between 82 brain regions measured through MRI in a sample of 28,159 individuals of European ancestry in the UK Biobank cohort.^20^ We focus on the analysis of shared (pleiotropic) genetic signal across brain connections and characterise the extent of genetic effects throughout the brain, allowing us to identify genes and variants with a central role in the genetics of FC. Specific locus-edge and gene-edge associations are using gene-set and pathway enrichment analysis to explore putative biological processes underlying the individual differences in specific brain circuitry. This study identifies pleiotropic genetic loci and biological pathways influencing functional brain connectivity, providing mechanistic insights into how genetic variation may influence heterogeneity in functional connectivity and brain-related disorders.

## Results

### Common genetic variants explain interindividual differences in the strength of functional connections in the human brain

We conducted 3,321 GWAS on functional connectivity between all brain connections across the human brain in a discovery sample of 24,442 subjects. These comprise all pairwise functional connections between 82 brain regions: 68 anatomically derived cortical areas and 14 subcortical nuclei (**Methods;** Overview of methods and results in **Suppl. Fig. 1**).

Median phenotypic correlation between edges was 0.002 (range = [-0.681; 0.953]).

SNP-heritability (h^2^_SNP_) was calculated with Linkage Disequilibrium Score Regression (LDSC: **Methods**)^21^ to estimate the global influence of common genetic variants (minor allele frequency > 1%) in all functional brain connections. Median estimated h^2^_SNP_ across all 3,321 edges was 3.63% and nominally significant effects were found in 1,083 edges (32.6% of total) with a median h^2^_SNP_ value of 6.2%. The most heritable edge had h^2^_SNP_ = 19.9% (SE = 2.5%) between the left supramarginal gyrus and left middle temporal gyrus, two brain regions involved in language processing (**Suppl. Table 1; Fig. 1a**).^22,23^ Heritability estimates of subcortical edges were on average lower (mean h^2^_SNP_ = 1.9%), likely due to the relatively low signal-to-noise ratio of FC between subcortical nuclei.^24^ These results indicate that a moderate amount of interindividual differences in FC can be explained by interindividual differences in common genetic variants.

Subnetwork enrichment analysis was performed to determine if brain connections within specific resting-state networks (RSNs) were under stronger genetic control. For this, a linear model was applied to aggregate the h^2^_SNP_ of individual edges and calculate RSN enrichment for h^2^_SNP_, while accounting for the higher expected correlation of edges belonging to the same network (**Methods**). RSNs were defined by assigning cortical edges to one of the 7 Yeo-Krienen RSNs or subcortical network if both regions each edge was linking belonged to the same RSN (**Suppl. Methods**). Enrichment testing was also performed for other edge categories (i.e. cortical left intrahemispheric, cortical right intrahemispheric, cortical interhemispheric, and subcortical edges; **Suppl. Note**). Out of all RSNs (median h^2^_SNP_ range = [1.9; 6.9]%), the Ventral Attention network was found to be the most significantly enriched for h^2^_SNP_ (median h^2^_SNP_ = 6.9%; t(269) = 2.27, p =.012; **Table 1**; **Fig. 1b)**, but not survived multiple testing correction.

**Figure 1.**
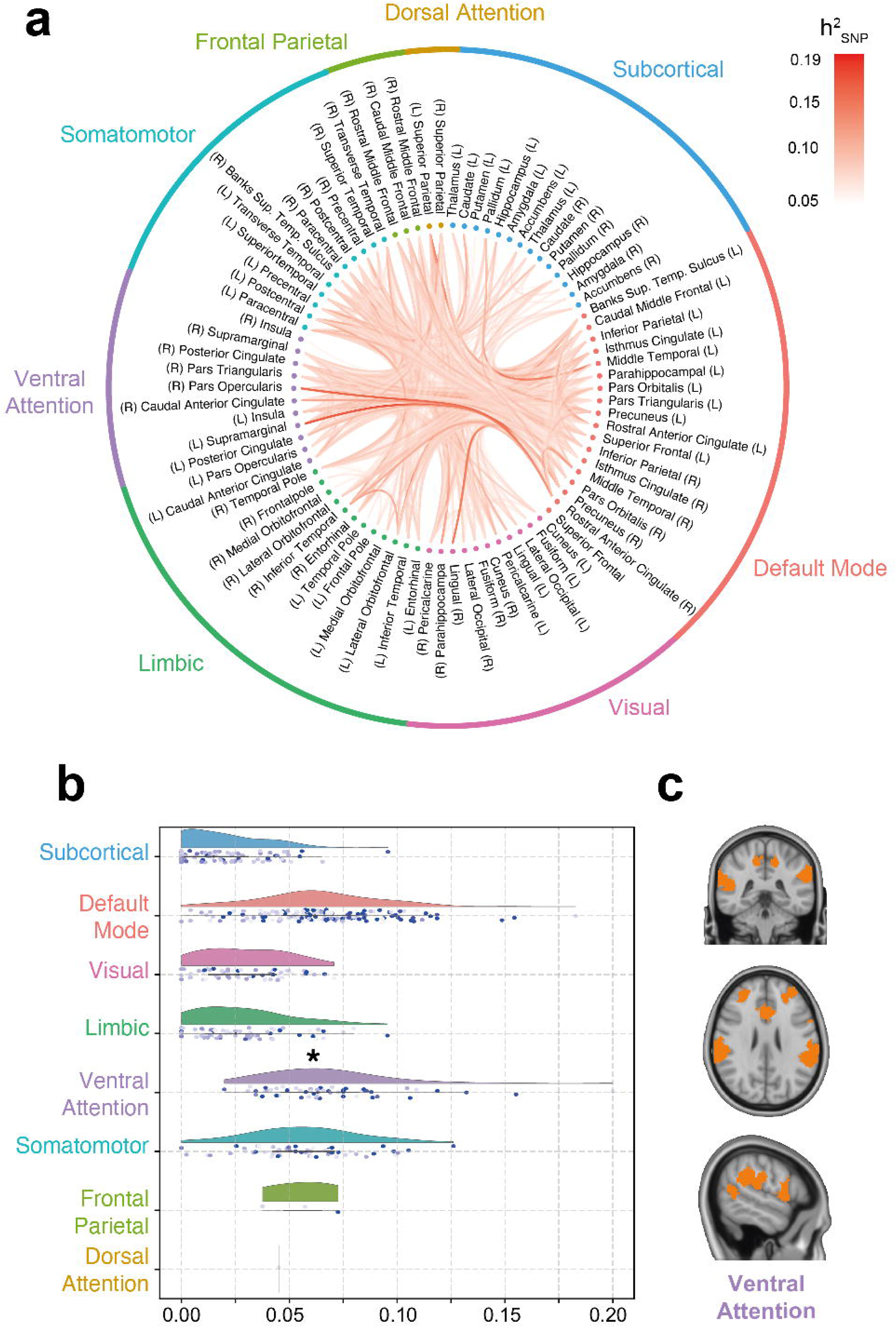
SNP-heritability of the human functional connectome. a | Circus plot of SNP-heritability estimates across the brain. b | Heritability estimates grouped per network. Half-violin plots represent the distribution of edge h^2^_SNP_. Each point below the violin plot is the heritability estimate for each edge belonging to the network. One asterisk (*) represents a p-value <.05 in subnetwork enrichment testing. The shade of points in the jitter plot depicts the standard deviation of the estimation with lighter points having more uncertainty. c | Visualisation of the Ventral Attention Network on the brain.

**Table 1.**
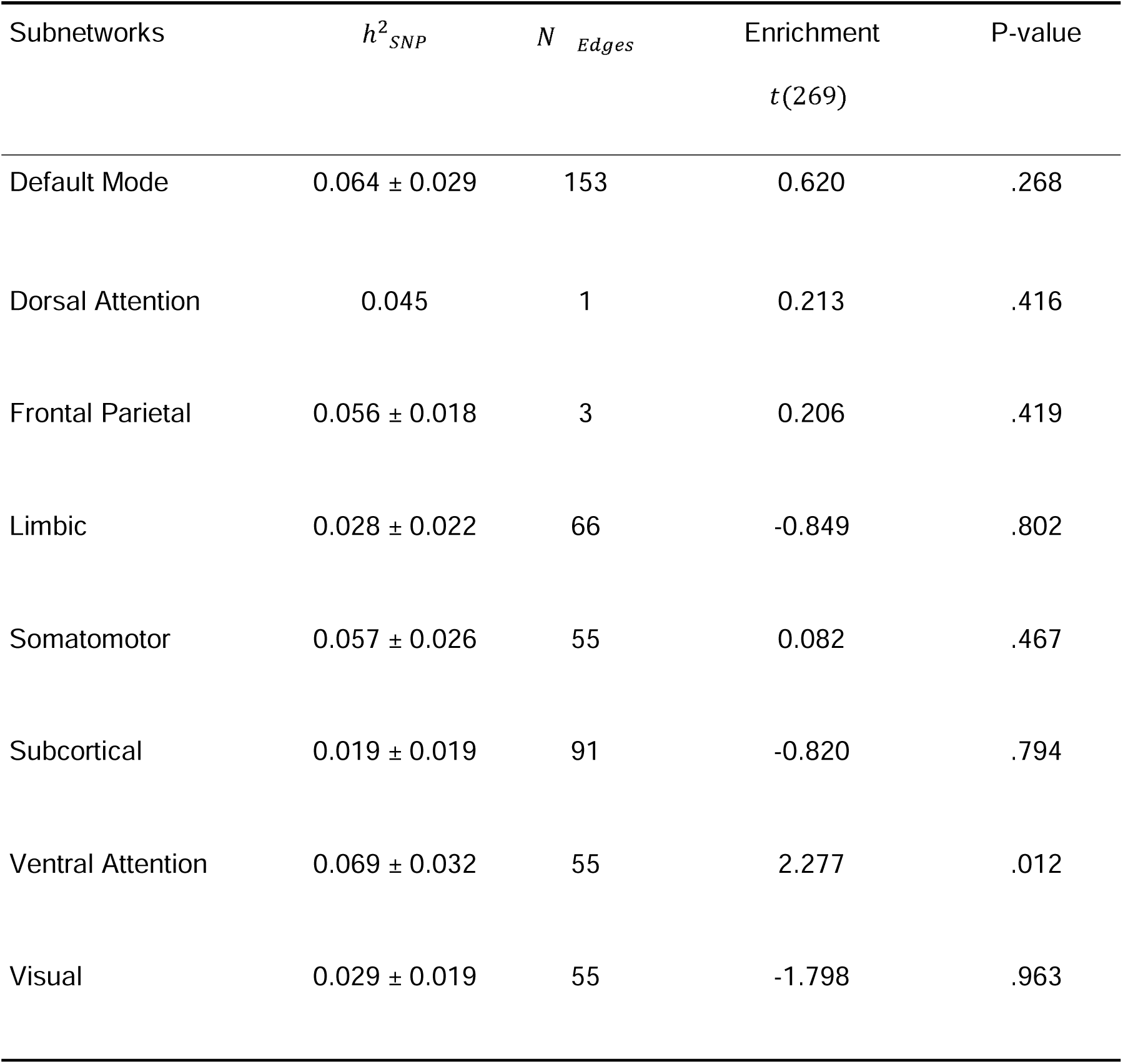
Enrichment for different subnetworks. h^2^_SNP_ represents the average LDSC SNP-heritability estimate of the edges in the subnetwork (mean ± standard deviation). N_edges_ is the number of edges in each subnetwork. Bonferroni-corrected significance is set at a one-sided a = 0.05/8.

### Seventy-eight replicated locus-edge associations within the human functional connectome

Loci in the genome associated with FC were identified in the discovery sample of 24,451 subjects and replicated in 3,708 subjects across 3,321 functional edges (**Methods**). Across all GWAS, a total of 208 significant locus-edge associations were identified in the discovery sample (α_disc_ =5 10^-8^/3321 for study-wide significance (SWS); **Methods**). Of those, 78 univariate locus-edge associations with 27 distinct edges were replicated in the holdout sample (α_disc_ = 0.05/208; **Suppl. Table 2**). SNP-heritability was found to be correlated with edgewise test-retest reliability, implying measurement noise affects the power to detect associated genetic variants (Spearman’s p = 0.68; **Suppl. Note**). Because of this, locus yield is likely to increase with increasing phenotype sample size.

The 78 SWS loci were often overlapping across edges or in close proximity in the genome. Locus-edge associations in the same genomic region were found to be co-localised in the discovery sample. This indicates that the same putative causal variants in these loci contribute to interindividual variability in multiple edges across the connectome (**Suppl. Note; Suppl. Table 3**). These locus-edge associations were aggregated into 4 non-overlapping genetic loci. To prioritise genes underlying the association of these loci with FC, effector gene prediction (EGP) with FLAMES was used.^25^ The 4 loci were mapped to genes *PAX8* (chromosome 2, base pairs: 113963070:114213070), *EphA3* (3:89451721:90010903), *THBS1* (15:39514832:39764832) and *APOE* (19:45286941:45536941; **Fig. 2a, top**; **Methods**; **Suppl. Table** 4, all positions in build GRCh37). The lead variant associated in the *APOE* locus was rs429358, a SNP coding for an exonic missense variant in the *APOE* gene (p.Cys130Arg), which is a well-known common risk variant for AD.^26^ The *PAX8* locus was associated at SWS level with 11 edges, *EphA3* with 15, *THBS1* with 4 and *APOE* with 1.

**Figure 2.**
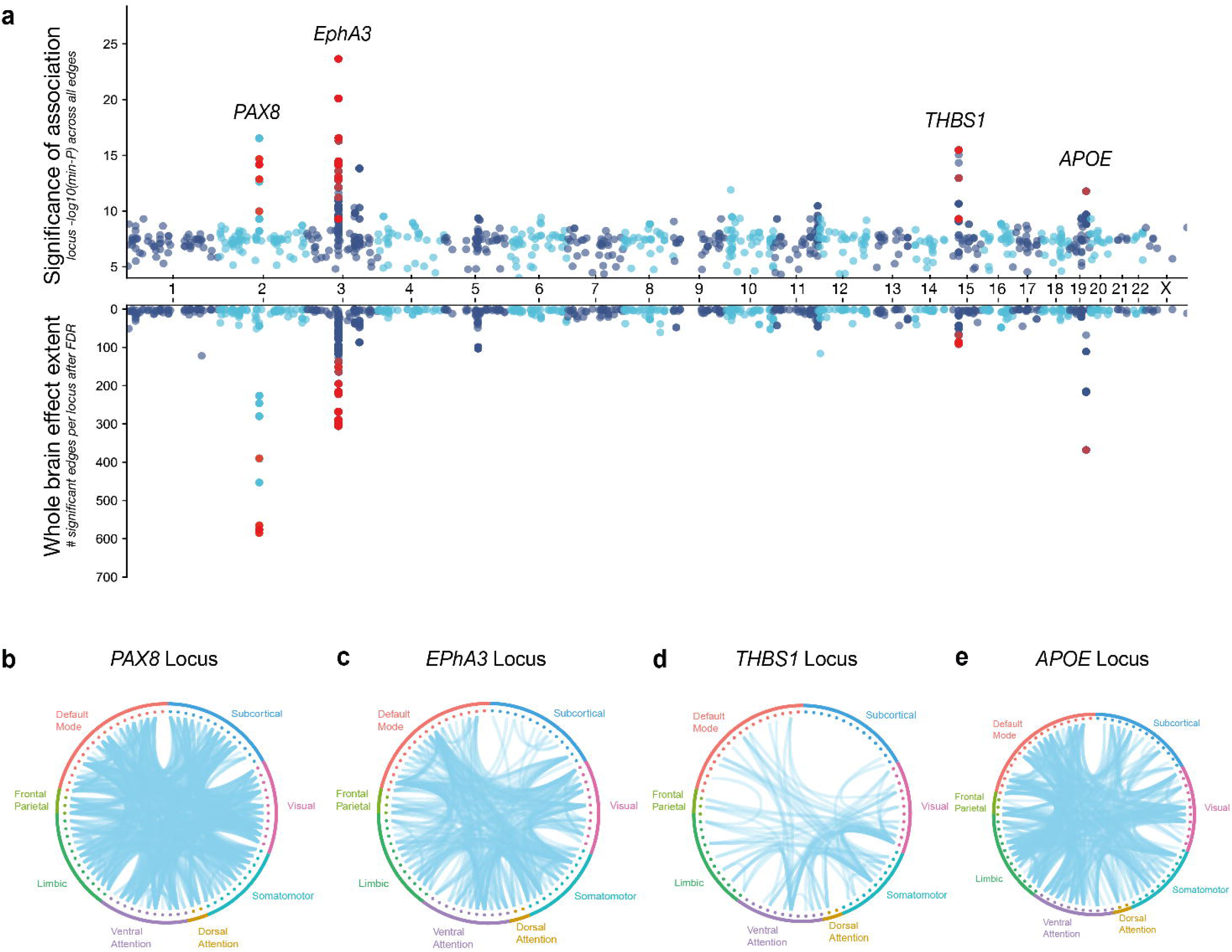
Pleiotropy of genome-wide significant locus-edge associations with functional connectivity. a | Miami plot for locus association and effect extent across 3,321 GWAS for edges. A locus for which the lead SNP was found to be significant in at least one edge-GWAS is represented by the dots (replicated locus-edge associations in red). Top: - log(min-P) of the association with the locus across all edges. Bottom: number of edges that pass FDR correction for locus significance (effect extent estimation). b-e | Circos plots of the connections predicted to be associated with each replicated locus. Only loci significant for an alpha level FDR-corrected for the number of phenotypes tested are drawn.

### Genetic loci relevant for functional connectivity are on average associated with 10% of all brain connections

The significance of these loci was compared across all functional edges to differentiate connectome-wide versus edge-specific effects in all genome-wide significant (GWS) loci. For this, significance in the *PAX8*, *EphA*3, *THBS1* and *APOE* loci were compared across all 3,321 phenotypes and a measure of the extent of the effect was calculated based on the relative significance of these loci across all edges (**Methods**). The median effect extent for the 4 replicated loci was 10% of all edges in the brain (337 edges out of 3,321; range: [92; 585]; **Suppl. Fig. 6**). Several other (non-replicated) GWS loci showed substantial pleiotropic effects across edges (**Fig. 2a, bottom**), but the effects of other loci in the brain are on average less widespread (median effect extent of all GWS loci = 0.18%; 6 edges out of 3,321 edges; range: [0%; 17%]). The *PAX8, APOE*, *EphA3* and *THBS1* loci were estimated to affect 17%, 11%, 9% and 3% of all functional edges in the brain (585, 368, 306 and 92 out of 3,321 edges), respectively. These results indicate these loci might have a widespread effect throughout the whole brain (i.e. are more pleiotropic), rather than having a very local effect (see **Fig. 2b-e** for spatial organisation of these associations**, Suppl. Fig. 2-5**).

### Functional connectivity shares a genetic basis with cardiovascular, cognitive and metabolic domains

The convergence of the SNP-based associations was additionally investigated using MAGMA to perform genome-wide gene-based association studies (GWGAS; **Methods**).^25,27,28^ A GWGAS was run per edge to combine individual SNP p-values into gene-edge association scores for a set of 18,852 protein-coding genes. GWGAS revealed associations with 6 unique genes, 4 of which had not been identified on the SNP-based analyses: *EphA3*, *APOE*, *APOC1*, *ZIC4*, *ZIC1* and *SLC39A12* (α_disc_ = 0.05/(18,852 3,321); **Suppl. Table 5**). Gene-edge associations with *EphA3* and *SLC39A12* were replicated in the holdout sample (**Methods**; **Suppl. Table 6**). *SLC39A12* is a novel association with FC between the left and right putamen. This gene has not previously been reported in FC GWAS and it is unique to GWGAS, being outside of the 4 loci discovered in SNP-based testing.^8,10,29–31^ Together with the results of EGP, 5 genes were found to be associated with FC: *EphA3*, with both GWGAS and EGP; *SLC39A12*, with GWGAS alone; *PAX8*, *THBS1* and *APOE*, with EGP alone.

Gene-set enrichment analysis (GSEA) was used to investigate whether genes associated with FC converge in specific biological processes and molecular functions, using hypergeometric testing in GENE2FUNC in FUMA.^27,28^ Two candidate gene-sets were analysed: a strict gene-set comprising the 5 genes with SWS replicated associations and a broad gene-set containing 322 different protein-coding genes associated at genome-wide significance level with at least one edge in GWGAS (α_GWS_ = 0.05/18,852; **Methods**; **Fig. 3a**). The strict candidate gene-set for FC was enriched for two Gene Ontology (GO) molecular functions (protein-lipid complex binding and proteoglycan binding) and nine GO biological processes (top three: tube morphogenesis, nitric oxide mediated signal transduction and regulation of cellular component biogenesis**; Suppl. Fig. 7; Suppl. Table 6**). Next, the second broader candidate gene-set was used to explore more general phenotypic associations using genes for which there is a considerable amount of subthreshold signal. While GWS genes form a bigger gene-set, the resultant gene-set is likely to be noisier. However, the broader set of genes was enriched for protein-protein interactions compared to what would be expected for a random set of proteins of the same size and degree distribution drawn from the genome (p = 1.03 · 10^-6^; **Suppl. Fig. 8**), which indicates that the proteins coded by these genes are biologically connected as a group.^32^ GSEA revealed fifty-three processes were significantly associated with FC. These were separated into 10 different domains, namely associations with cardiovascular (9 processes), cognitive (9), metabolic (7), longevity (5), imaging-derived (5), alcohol consumption (4), dietary (3), neurological (3), gastrointestinal (2) and respiratory (2) domains (**Fig. 4**; **Suppl. Fig. 9**; **Suppl. Table 8**). No GO terms, tissue or cell types, or curated gene-sets were significantly enriched for the broad gene-set.

**Figure 3.**
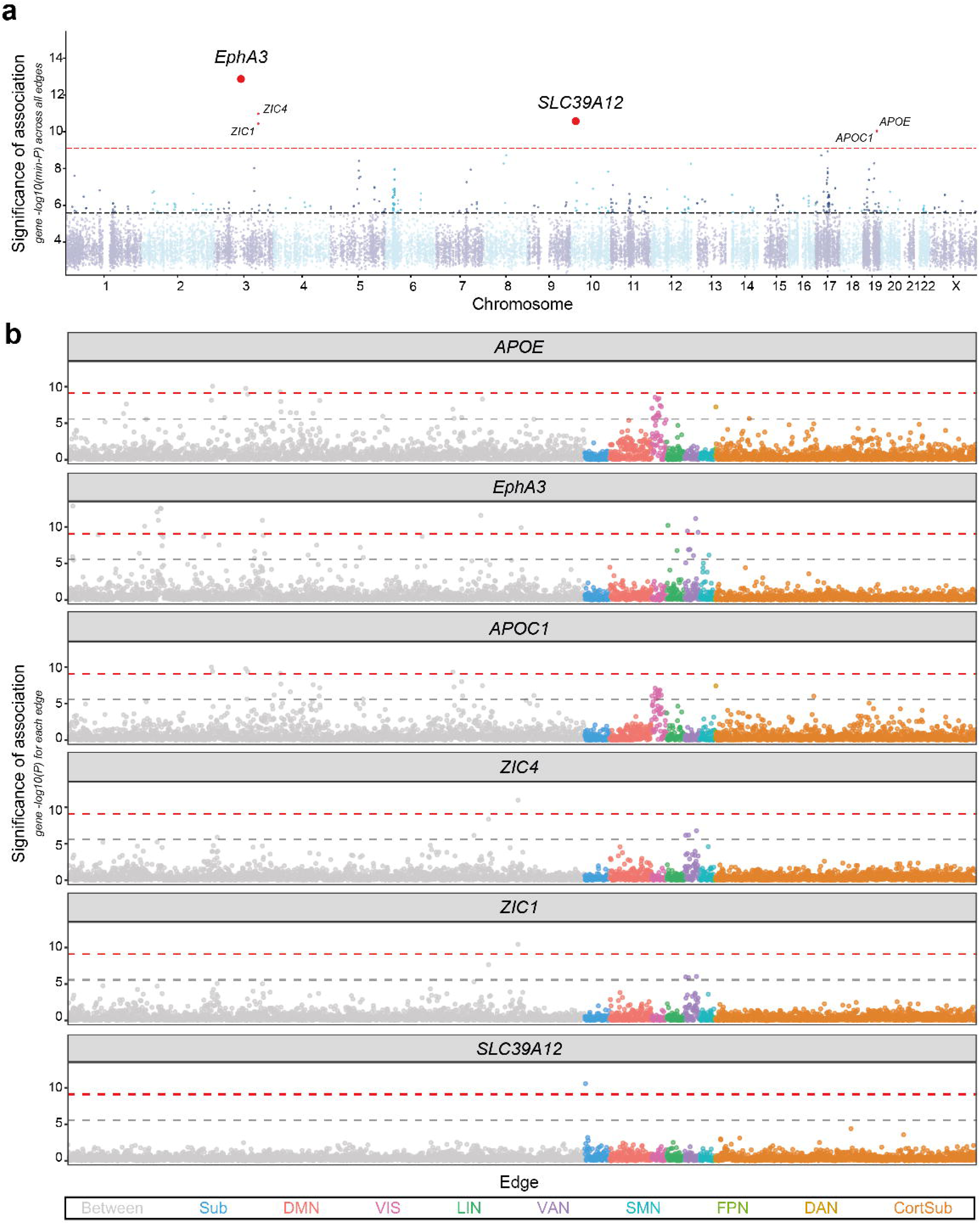
Gene-based testing functional connectivity. a | Manhattan plot for GWGAS across 3,321 functional brain connections. Each dot represents one protein-coding gene. The height of the point depicts the-log(min-P) of the association with all the phenotypes, i.e. the strongest association across all phenotypes. Genes with a min-P value significant after correcting for the number of genes and phenotypes are in red. EphA3 and SLC39A12 were replicated in a holdout sample. b | Univariate gene-edge association p-values for the 6 SWS genes discovered through GWGAS. From those *SLC39A12* and *APOE* were replicated. Each point represents an edge and the colour of the point the subnetwork it belongs to. Between: Edge between networks. Sub: Subcortical. DMN: Default Mode Network. VIS: Visual Network. LIN: Limbic Network. VAN: Ventral Attention Network. SMN: Somatomotor Network. FPN: Frontoparietal Network. DAN: Dorsal Attention Network. CortSub: Cortico-subcortical edge. Dashed lines represent study-wide significance (α_SWS_ = 0.05/(18,852 · 3,321), in red) and genome-wide significance (α_GWS_ = 0.05/18,852, in grey) for gene-based testing.

**Figure 4.**
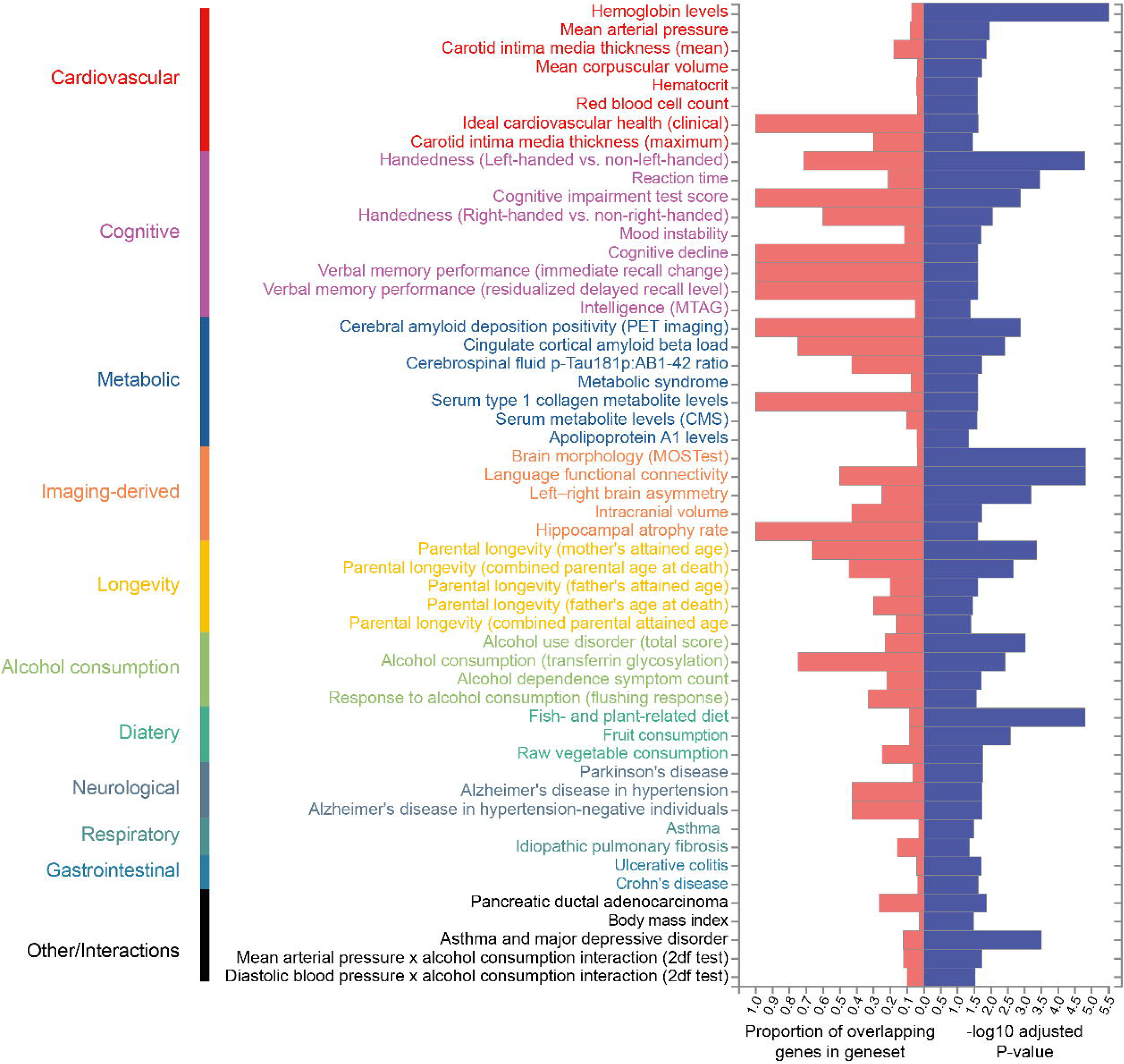
Gene-set enrichment analysis for all annotated genes (N genes = 322). Significantly enriched gene-sets were manually sorted by domain.

We further used the effect extent definition to determine whether genes involved in neuropsychiatric disorders are expected to be associated with more brain functional connections than an equivalent set of highly brain-expressed genes (**Suppl. Methods**; **Suppl. Note**). When comparing the effect extent of gene-sets consisting of significant genes for several neuropsychiatric disorders, a significantly larger extent was found for genes linked to attention-deficit/hyperactivity disorder (ADHD; p =.04),^33^ anorexia nervosa (p =.03),^34^ Alzheimer’s disease (AD; p <.0001),^35^ bipolar disorder (p =.02)^36^ and schizophrenia (p <.0001).^37^ After correcting for multiple testing (α = 0.05/6, for 6 disorders tested), the gene-sets for AD and schizophrenia remained significant (**Suppl. Fig. 10**). These results suggest that genes associated with neuropsychiatric disorders are expected to have a widespread effect in FC, implying a common genetic basis for alterations in FC and neuropsychiatric diseases.

### Functional interpretation of gene-edge associations points towards neurodevelopmental and angiogenic processes in the genetic control of the functional connectome

The most significant locus-edge association was linked to the *EphA3* gene (min-p_locus_ = 2.19·10^-24^). This gene is among the top 10% of human genes most intolerant to variation, meaning that mutations are likely to lead to downstream biological consequences.^38^ The receptors coded by *EphA3* are involved in neurodevelopmental events such as lobe-specific axonal guidance and cortical expansion of the primate brain.^39–41^ The *EphA3* gene has been previously associated in prior GWAS of language-related FC phenotypes and structural connectivity.^29,42^ This work extends the prior association of this gene with structural connectivity beyond functional language circuits, showing a widespread association with FC.

The most pleiotropic association was linked to the *PAX8* gene in chromosome 2 (min-p_locus_ = 2.92·10^-17^, effect extent = 17% of all edges). This master regulator transcription factor gene has been associated with several FC phenotypes, from graph measures^30^ to RSNs^8^ and individual edges in the UK Biobank cohort.^29^ It has been related to endocrine tissues, and shown to play a key role in epithelial cell survival and proliferation.^43–45^ This genetic signal associated with this gene co-localised within all edges with which it was found to be associated, implying a highly localised genetic influence on FC.

The *APOE* and *THBS1* genes were found to drive the enrichment of FC for biological processes of proteoglycan binding (enrichment-p = 3.57·10^-5^) and protein-lipid complex binding (enrichment-p = 2.0·10^-5^; **Suppl. Fig. 7**; **Suppl. Table 7**). These have both been reported to be genetic underpinnings for AD.^35,46–48^ The *APOE* gene codes for apolipoprotein E, which is mostly synthesised by microglia and astrocytes in the brain and plays a fundamental role in cholesterol metabolism.^47^ It has been consistently related to longevity^47^ and heart health^49^, phenotypes found to be genetically associated with FC in our study (**Fig. 3b**). *THBS1* is a gene coding for thrombospondin-1, a glycoprotein found in the extracellular matrix involved in postnatal neuronal migration.^50^ We found enrichment for several biological processes related to angiogenesis (e.g. tube morphogenesis and development, negative regulation of blood vessel endothelial cell migration and regulation of sprouting angiogenesis; see **Suppl. Fig. 5** and **Suppl. Table 7**). The lifetime expression of these genes is different: *APOE* is highly expressed in the brain postnatally until late adulthood, whereas the peak expression of *THBS1* is perinatal (**Suppl. Fig. 11,12**). The *APOE* gene has also been specifically associated with changes in brain morphology across the lifespan.^13^ Our findings indicate a diverse genetic modulation of FC throughout the lifetime, with a neurodevelopmental and an older age component.

A novel gene-edge association was found between *SLC39A12* and FC in the bilateral putamen (min-p_gene_ = 2.70·10^-11^; **Suppl. Table 6**). This gene is differentially expressed in astrocytes and the choroid plexus.^51^ It is involved in the transportation of zinc from the extracellular space to the cytoplasm across the cell membrane, having been implicated in the processes of neurulation and neurite extension and shown to have an altered expression profile in schizophrenia.^52,53^ *SLC39A12* has previously been associated with anatomic alterations in the putamen, among other cortical and subcortical brain regions (T1 and T2*-imaging).^9,12,18^ These changes were hypothesised to be associated with iron deposition in ageing and pathology.^12^ Rare predicted loss-of-function variants in this gene have no other reported associations with associations with imaging or neuropsychiatric phenotypes.^54^ These results, together with our novel association with FC, suggest that S*LC39A12* might underlie both imaging and psychiatric phenotypes.

## Discussion

Mechanistic insight into how different brain areas are connected is essential to understanding the functioning of the brain. The biological processes underlying these circuits support the maintenance of an efficient information flow across the brain, with importance in health and disease.^55^ We investigated the biological underpinnings of the human functional connectome by parcellation-based GWAS analysis of connection-wise functional brain connectivity. Our findings show a complex landscape of genetic influences on functional brain networks and illustrate the importance of genetic heterogeneity for differences in resting-state brain activity across individuals. Genetic associations found independently for different functional connections converge into a genetic basis shared with both physiological and cognitive traits, placing the functional connectome in the intersection between brain and physical health.

Family studies^6,56^ and GWAS^8,31^ have reported resting-state networks (RSNs) to be heritable, but the heritability of all functional links of the human connectome together remained unknown. We find that over 30% of connections in the human functional connectome are significantly heritable, for which all common (MAF>1%) variants in the genome explain on average 6.5% of interindividual variation. This number is comparable to the 9.1% found for anatomical connectivity using comparable methods.^42^

Four genes — *PAX8*, *EphA3*, *THBS1*, and *APOE* — were found to particularly stand out. These genes have been previously shown to influence macroscale brain connectivity, with *EphA3* playing a role in both anatomical and functional connectivity.^8,11,29,42,57^ This supports the idea of high pleiotropy between MRI-derived measures and neuropsychiatric traits ^17,19,58,59^ Despite their known genetic associations with various FC measures,^8,9,29^ *APOE*, *PAX8*, and *THBS1* were not linked to anatomical edge-level connectivity in recent large-scale GWAS.^11,42^

In addition to the four genes above, we found an association with *SLC39A12*. To our knowledge, this is the first time *SLC39A12* has been linked to brain connectivity, highlighting its potential as a new genetic factor influencing FC. *SLC39A12* encodes a zinc transporter involved in metal ion homeostasis, which has been implicated in neurodevelopment and synaptic function.^52,53^ This finding suggests that distinct molecular mechanisms may underlie different aspects of brain organisation measured through MRI.

Functional annotation of the set of putative FC genes revealed associations with cardiovascular, cognitive and metabolic phenotypes. Cardiovascular health has been shown to play an important role in the maintenance of FC, both phenotypically^60–62^ and genetically.^57^ We find cognitive phenotypes to be genetically associated with FC, connecting the genetic factors of FC and those of cognition. In the literature, GWAS on brain networks specific to schizophrenia-and bipolar disorder-specific brain networks showed that risk for these disorders and connectomic alterations share associations with the Notch signalling pathway, which is involved in functions of neural development and blood vessel formation, among other processes.^63^ This suggests a role of cardiovascular health and FC in supporting healthy cognition, with angiogenesis being a putative mechanism for the relationship between these three domains. Moreover, enrichment for phenotypes in the metabolic domain, together with the association with *APOE*, suggests a link between the genetic factors influencing FC and AD. In particular, genetic associations between FC and cerebrospinal fluid p-Tau181p:AB1-42 ratio, cerebral amyloid deposition positivity, apolipoprotein A1 levels and cingulate cortical amyloid beta load point towards important processes in the pathophysiology of AD.^64–66^ These findings highlight extensive pleiotropy between brain connectivity and traits from several phenotypic domains, indicating an interplay between cognition, functional brain connectivity and physical health on a genetic level.

This study has important limitations to consider when interpreting its findings. First, a mass-univariate method entails a significant loss of power due to the multiple-testing burden. Therefore, study designs such as performed here are mostly attuned to finding large pleiotropic results, despite the use of a mass-univariate method. Estimates of locus yield, effect extents, SNP-heritability should be considered within the current power level and the differential power to detect genetic effects across the brain. Secondly, locus effect extent calculations do not consider the correlational structure of the connectome. These should be interpreted relative to one another rather than as an absolute value. Next, the use of a single cohort of individuals of older adults with European ancestry may limit the generalisability of our findings, particularly considering the extensive literature in imaging-genetics using the UKB as a study cohort. Sample-specific characteristics like volunteering and ascertainment biases, older age range, and socioeconomic status of subjects may also introduce unexpected stratification effects which may partially confound genetics and environmental effects. Finally, the small size of the strict candidate gene-set makes GSEA sensitive to chance effects. The statistical significance of the highlighted biological processes should be interpreted with caution.

In this work, we contextualise the genetic basis of the human functional connectome in relation to physical and neurocognitive traits. Our findings show that brain connectivity and phenotypes from physical and cognitive domains share a genetic basis, extending the pleiotropy found across the functional brain connections to other dimensions of physical and cognitive health.

## Data Availability

Genome-wide summary statistics will be made available upon publication at https://cncr.nl/research/summary_statistics/.
LD reference for LDSC, https://www.internationalgenome.org/category/reference/; RSN Annotation Files, https://surfer.nmr.mgh.harvard.edu/fswiki/CorticalParcellation_Yeo2011. FLAMES pathway-naive feature set: https://zenodo.org/records/10409723; Case-control disease sumstats: https://pgc.unc.edu/for-researchers/download-results/.

## Acknowledgements

D.P., C.R., B.A.P.C.M, M.P.vd.H., and M.S. were funded by NWO Gravitation: BRAINSCAPES: A Roadmap from Neurogenetics to Neurobiology (grant no. 024.004.012 [to D.P.]). D.P. and C.d.L. were funded by a European Research Council advanced grant (no. ERC-2018-AdG GWAS2FUNC 834057 [to D.P.]). J.E.S. is supported by the NWO VENI Grant 201G-064. This manuscript was submitted to bioRxiv. This research has been conducted using the UK Biobank resource under application 16406. We thank the numerous participants, researchers, and staff who collected and contributed to the data. We thank SURF (www.surf.nl) for the support in using the National Supercomputers Snellius and LISA. We thank Dr Rachel M. Brouwer, Dr Sophie van der Sluis and Ilan Libedinsky for all the feedback. Finally, we would also like to acknowledge Dr Elleke P. Tissink and Dr Siemon C. de Lange, who preprocessed the MRI data.

## Author Contributions

B.d.A.P.C.M.: Conceptualisation, Methodology, Formal Analysis, Writing, Visualisation. M.S.: Formal Analysis, Feedback. C.R.: Visualisation, Feedback. C.d.L: Methodology, Feedback. K.H.: Feedback. D.P.: Funding acquisition, Feedback. J.E.S.: Methodology, Supervision, Feedback. M.P.v.d.H: Conceptualisation, Methodology, Resources, Supervision, Project Administration.

## Online Methods

### Study Cohort

Genetic and neuroimaging data from the UK Biobank (UKB), a large-scale cohort with neuroimaging, genotyping, clinical, demographic, and behavioural data of volunteer participants from a population-based sample of older adults in the UK were used in the present study.^20^ The UKB initiative has been approved by the National Research Ethics Service Committee (reference 11/NW/0382) and data were accessed under application number 16406. Joint neuroimaging and genotypic data (imputed from Single Nucleotide Polymorphism (SNP) arrays) were available for a total of 40,682 subjects. From those, 5,000 subjects were randomly assigned to a holdout sample before any analysis for replication purposes. Subjects with sex aneuploidy, discordant reported and chromosomal sex, high degrees of relatedness (n = 5,241) and non-European (n = 2,034) individuals were excluded, as previously described.^67^ After quality control, a total of 24,451 subjects in the discovery sample and 3,708 in the holdout sample were included in our analysis. A detailed overview of exclusions and a full description of the demographic characteristics of the discovery and holdout samples are provided in **Suppl. Note**.

### Neuroimaging and Genotypic Data Preprocessing

UK Biobank imputed genotyping SNP-array data were used for this study (in build GRCh37). The imputation and quality control performed by the UK Biobank team are described extensively elsewhere.^20^ After additional in-house genomic quality control (**Suppl. Methods**), a total of 9,380,668 high-quality SNPs were hard-called under a certainty threshold of 0.9.^68^ The acquisition and preprocessing protocols for the neuroimaging data are described in the UKB Brain Imaging documentation.^69^ Resting-state functional magnetic imaging (rs-fMRI) was used with T1 surface model files and structural segmentation from FreeSurfer (v6.0). Functional connectivity was computed using CATO (Connectivity Analysis Toolbox; v3.1.6).^70^ Coregistration was performed by aligning the subject average of rs-fMRI across all time points with the T1 image.^71^ Next, surface-based cortical parcellation was performed using FreeSurfer parcellating the cortical mantle according to the Desikan-Kiliany atlas.^72^ Subcortical volumes were automatically segmented into the aseg atlas in the T1 image.^73^ This resulted in a whole brain parcellation of a total of 82 brain areas (68 cortical + 14 subcortical). Motion metrics, alongside their first-order drifts and their linear trends, and the average signal of voxels in both cerebrospinal fluid and white matter were regressed out of the rs-fMRI signal. A zero-lag bandpass filter ([0.01-0.1] Hz band) and motion-scrubbing were applied to the time series: frames with more than 2 violations were discarded alongside with their closest backward neighbour (maximum framewise displacement = 0.25, maximum DVARS = 1.5).^74^ FC matrices were constructed by calculating pairwise Pearson’s correlations between the average preprocessed time series of each of the 82 parcellated brain areas. Subjects for which average head motion parameters (across space and time), signal-to-noise ratio or discrepancy between T1-weighted and fMRI scan deviated from the median more than five times their median absolute variation were excluded from the analysis.^75^

### SNP-based GWAS

Identification of common genetic variants involved in FC was carried out using PLINK2.0.^76^ A total of 3,321 SNP-based GWAS on single-edge connectomic data were performed for 8,790,386 imputed and genotyped SNPs. Phenotypes were standardised across subjects and residualised for total intracranial volume. This analysis was performed on independent (Linkage Disequilibrium threshold, r^2^ > 0.1), common (MAF > 0.1), and genotyped SNPs or SNPs with very high imputation quality (INFD > 0.9). The first 20 principal components (extracted using FlashPCA2;^77^ **Suppl. Note**) were used as covariates in all GWAS together with sex, age, handedness, genotype array, and covariates specific to fMRI: scanning site, time to echo, table coordinates, coil position, signal-to-noise ratio, mean head motion, intensity scaling parameters and framewise displacement.^75^ The full set of covariates was standardised. Male X variants were coded as 0/1 (assuming no deactivation) to avoid double counting.^78^

### SNP-heritability estimation

Using linkage disequilibrium score regression (LDSC; v.1.0.1),^21^ SNP-based heritability for each edge-GWAS was estimated. After munging,^21^ SNPs in the HAPMAP3 reference panel were included (**Data Availability**), leaving 1,137,297 SNPs for analysis. The significance level for heritability was defined as α_h_2 = 0.05. Additionally, a cut-off λ > 1.02 was set to determine which phenotypes had enough polygenic signal to run LDSC and for the LDSC intercept (Intercept < 1.1) to control confounding from population stratification.^21^

A linear regression model with a fixed variance-covariance structure of the residuals was used to determine whether any of the 7 Yeo-Krienen RSNs^1^ (and subcortical network) was enriched for the h^2^_SNP_. The genetic covariance matrix was estimated through the calculation of edge-wise correlations between each pair of edges for all the individuals allocated to the discovery sample. The significance level was set at α_subnetwork_ = 0.05/8 to correct for multiple testing for the 7 RSNs and subcortical network. See **Suppl. Methods** for more details on the method.

### Definition and effect extent of genomic risk loci

The identification of relevant independent genomic loci was done using the linkage disequilibrium-based result clumping procedure implemented in PLINK1.9.^76^ Independent significant genome-wide (GWS) SNPs were defined as those with p < 5 · 10^-8^ for each trait in our discovery sample. An additional multiple testing correction was applied to account for the number of traits: if SNPs associated with a p < 5 · 10^-8^/3,321, they were considered study-wide significant (SWS). The 1,000 Genomes European subset was used as an LD reference for our analysis (**Data Availability**).^79^ Then, we clumped together all of the variants with p < 0.01 that are in LD with (LD threshold, r^2^ > 0.9) and in a radius of 250kb from each independent significant SNP. The most significant SNP (i.e., lowest p-value) is referred to as the ‘lead SNP’ of the locus. A locus-edge association refers to each [edge; lead SNP] pair for which the lead SNP was found to be genome-wide significantly associated with the functional connection. Only SWS loci were mapped onto genes.

A total of 1490 locus-edge associations (864 unique lead SNPs) were discovered at GWS level and further assessed across all brain connections to estimate the effect extent of these loci throughout the brain. For each of the 864 loci defined by unique lead SNPs, the p-values for all SNPs were combined using an SNP-wise mean model in MAGMA (multimarker analysis of genomic annotation; v1.10),^28^ resulting in one p-value per locus for each edge. Benjamini-Hochberg FDR correction for the number of traits (3,321) was applied across edges to these p-values to quantify with how many traits each locus was expected to be associated. This number is referred to as the effect extent of a locus. See **Suppl. Method** for extended methodology.

### Genome-Wide Gene Association Studies

Genome-Wide Gene Association Studies (GWGAS) was used to detect (whole) gene-edge associations in the functional connectome. Each SNP-based GWAS summary statistics was used to perform a GWGAS in MAGMA.^28^ A SNP-wise mean model was applied to the summary statistics using a reference of 10K individuals from our sample to test the jointassociation of all SNPs within 18,852 protein-coding genes with each trait. A multiple-testing corrected significance threshold α_gene_ = 0.05/(18,852 · 3,321) was used.

### Replication

Replication of the discovered locus-edge and gene-edge associations was performed in the hold-out sample of 3,708 individuals. A locus-edge association was considered replicated if the lead SNP in that locus for that edge (i) had a concordant effect direction in the discovery and replication samples, (ii) had MAF > 0.1 in the replication sample, and (iii) was Bonferroni-significant at α_SNP,rep_ = 0.05/208 (208 SWS locus-edge associations discovered). A gene-edge association was considered replicated if the gene was Bonferroni-significant for that edge in the replication sample at a a_gene,rep_ = 0.05/6 (6 significant genes tested).

### Prioritisation of genes and functional annotation

Two loci from two different GWAS were considered to overlap if their respective lead SNPs were separated by less than 250K base pairs. These loci were validated by co-localising the different GWAS with coloc (v5.2.2; **Suppl. Note, Suppl. Table 3**).^80^ Estimation of the LD reference panel consisting of all the individuals in each discovery GWAS was carried out on LDStore2 (v2.0).^81^ The resulting LD reference panel was used to fine-map each of the overlapping loci with FINEMAP (v1.4.1),^82^ using a threshold of a maximum of 10 causal variants per locus. The effector gene prediction (EGP) for each credible set was carried out using FLAMES (Fine-mapped Locus Assessment Model of Effector geneS; v1.0.0) with a 750 kb window on each side of the centroid of the credible set and prediction of coding genes.^25^ The Polygenic Priority Scores (PoPS; v0.2)^83^ for MAGMA were calculated using the pathway-naïve feature set provided by the authors (**Data Availability**). Predictions with a FLAMES score above 0.05 were kept for further analyses.^25^

Pathway convergence of the gene-sets resulting from univariate gene-based testing was investigated using GENE2FUNC in FUMA (Functional Mapping and Annotation of Genome-Wide Association Studies).^27^ GENE2FUNC performs hypergeometric tests to test if genes of interest are overrepresented in any of the pre-defined gene sets. A broad gene-set containing all associations across all GWGAS + EGP (322 genes) and the gene-set of replicated genes (1 + 4 genes) were tested separately. Enrichment testing was performed using a background of all protein-coding genes. Pathway-specific enrichment was calculated for gene-sets from Gene Ontology molecular functions, cellular components and biological processes, and curated gene-sets from MsigDB (gene-set categories C2 and C5; v7.0) and all gene-sets in the GWAS catalog.^84,85^ Gene-sets where at least two of our input genes featured were considered. Benjamini-Hochberg multiple testing correction per gene-set category for the hypergeometric gene-set enrichment testing and tissue differential expression analysis was adopted.

### Data availability

Genome-wide summary statistics will be made available upon publication at https://cncr.nl/research/summary_statistics/.

LD reference for LDSC, https://www.internationalgenome.org/category/reference/; RSN Annotation Files, https://surfer.nmr.mgh.harvard.edu/fswiki/CorticalParcellation_Yeo2011. FLAMES pathway-naïve feature set: https://zenodo.org/records/10409723; Case-control disease sumstats: https://pgc.unc.edu/for-researchers/download-results/.

### Code availability

All used software tools and code are publicly available: CATO, http://www.dutchconnectomelab.nl/CATO/; FUMA, http://fuma.ctglab.nl/; MAGMA, https://ctg.cncr.nl/software/magma; LDSC, https://github.com/bulik/ldsc; PLINK, https://www.cog-genomics.org/plink/; FlashPCA2, https://github.com/gabraham/flashpca; FLAMES, https://github.com/Marijn-Schipper/FLAMES; FINEMAP & LDStore2, http://www.christianbenner.com/; coloc R package, https://chr1swallace.github.io/coloc/.

## References

1. Thomas Yeo, B. T., et al. The organization of the human cerebral cortex estimated by intrinsic functional connectivity. J. Neurophysiol. 106, 1125–1165 (2011).

2. Petersen, S. E. & Sporns, O. Brain Networks and Cognitive Architectures. Neuron 88, 207–219 (2015).

3. van den Heuvel, M. P. & Sporns, O. A cross-disorder connectome landscape of brain dysconnectivity. Nat. Rev. Neurosci. 20, 435–446 (2019).

4. Fornito, A., Zalesky, A. & Breakspear, M. The connectomics of brain disorders. Nat. Rev. Neurosci. 16, 159–172 (2015).

5. Gu, Z., Jamison, K. W., Sabuncu, M. R. & Kuceyeski, A. Heritability and interindividual variability of regional structure-function coupling. Nat. Commun. 12, 4894 (2021).

6. Glahn, D. C. et al. Genetic control over the resting brain. Proc. Natl. Acad. Sci. 107, 1223–1228 (2010).

7. Reineberg, A. E., Hatoum, A. S., Hewitt, J. K., Banich, M. T. & Friedman, N. P. Genetic and Environmental Influence on the Human Functional Connectome. Cereb. Cortex 30, 2099–2113 (2020).

8. Tissink, E. et al. The Genetic Architectures of Functional and Structural Connectivity Properties within Cerebral Resting-State Networks. eNeuro 10, (2023).

9. Elliott, L. T. et al. Genome-wide association studies of brain imaging phenotypes in UK Biobank. Nature 562, 210–216 (2018).

10. Zhao, B. et al. Common variants contribute to intrinsic human brain functional networks. Nat. Genet. 54, 508–517 (2022).

11. Sha, Z., Schijven, D., Fisher, S. E. & Francks, C. Genetic architecture of the white matter connectome of the human brain. Sci. Adv. 9, eadd2870 (2023).

12. Smith, S. M. et al. An expanded set of genome-wide association studies of brain imaging phenotypes in UK Biobank. Nat. Neurosci. 24, 737–745 (2021).

13. Brouwer, R. M. et al. Genetic variants associated with longitudinal changes in brain structure across the lifespan. Nat. Neurosci. 25, 421–432 (2022).

14. Régy, M. et al. The role of dementia in the association between APOE4 and all-cause mortality: pooled analyses of two population-based cohort studies. Lancet Healthy Longev. 5, e422–e430 (2024).

15. Romero, C. et al. Exploring the genetic overlap between twelve psychiatric disorders. Nat. Genet. 54, 1795–1802 (2022).

16. Grotzinger, A. D. et al. Genetic architecture of 11 major psychiatric disorders at biobehavioral, functional genomic and molecular genetic levels of analysis. Nat. Genet. 54, 548–559 (2022).

17. Roelfs, D. et al. Shared genetic architecture between mental health and the brain functional connectome in the UK Biobank. BMC Psychiatry 23, 461 (2023).

18. van der Meer, D. et al. Understanding the genetic determinants of the brain with MOSTest. Nat. Commun. 11, 3512 (2020).

19. Tissink, E. P. et al. Abundant pleiotropy across neuroimaging modalities identified through a multivariate genome-wide association study. Nat. Commun. 15, 2655 (2024).

20. Bycroft, C. et al. The UK Biobank resource with deep phenotyping and genomic data. Nature 562, 203–209 (2018).

21. Bulik-Sullivan, B. K. et al. LD Score regression distinguishes confounding from polygenicity in genome-wide association studies. Nat. Genet. 47, 291–295 (2015).

22. Oberhuber, M. et al. Four Functionally Distinct Regions in the Left Supramarginal Gyrus Support Word Processing. Cereb. Cortex N. Y. NY 26, 4212–4226 (2016).

23. Davey, J. et al. Exploring the role of the posterior middle temporal gyrus in semantic cognition: Integration of anterior temporal lobe with executive processes. Neuroimage 137, 165–177 (2016).

24. Seoane, S., Modroño, C., González-Mora, J. L. & Janssen, N. Medial temporal lobe contributions to resting-state networks. Brain Struct. Funct. 227, 995–1012 (2022).

25. Schipper, M. et al. A framework for accurate prediction of effector genes in genetic loci for complex traits. 2023.12.23.23300360 Preprint at 10.1101/2023.12.23.23300360 (2023).

26. Fernández-Calle, R. et al. APOE in the bullseye of neurodegenerative diseases: impact of the APOE genotype in Alzheimer’s disease pathology and brain diseases. Mol. Neurodegener. 17, 62 (2022).

27. Watanabe, K., Taskesen, E., van Bochoven, A. & Posthuma, D. Functional mapping and annotation of genetic associations with FUMA. Nat. Commun. 8, 1826 (2017).

28. de Leeuw, C. A., Mooij, J. M., Heskes, T. & Posthuma, D. MAGMA: Generalized Gene-Set Analysis of GWAS Data. PLoS Comput. Biol. 11, e1004219 (2015).

29. Mekki, Y. et al. The genetic architecture of language functional connectivity. NeuroImage 249, 118795 (2022).

30. Foo, H. et al. Novel genetic variants associated with brain functional networks in 18,445 adults from the UK Biobank. Sci. Rep. 11, 14633 (2021).

31. Zhao, B. et al. Genetic influences on the intrinsic and extrinsic functional organizations of the cerebral cortex. 2021.07.27.21261187 Preprint at 10.1101/2021.07.27.21261187 (2022).

32. Szklarczyk, D. et al. The STRING database in 2023: protein-protein association networks and functional enrichment analyses for any sequenced genome of interest. Nucleic Acids Res. 51, D638–D646 (2023).

33. Demontis, D. et al. Genome-wide analyses of ADHD identify 27 risk loci, refine the genetic architecture and implicate several cognitive domains. Nat. Genet. 55, 198–208 (2023).

34. Watson, H. J. et al. Genome-wide association study identifies eight risk loci and implicates metabo-psychiatric origins for anorexia nervosa. Nat. Genet. 51, 1207–1214 (2019).

35. Wightman, D. P. et al. A genome-wide association study with 1,126,563 individuals identifies new risk loci for Alzheimer’s disease. Nat. Genet. 53, 1276–1282 (2021).

36. Mullins, N. et al. Genome-wide association study of more than 40,000 bipolar disorder cases provides new insights into the underlying biology. Nat. Genet. 53, 817–829 (2021).

37. Trubetskoy, V. et al. Mapping genomic loci implicates genes and synaptic biology in schizophrenia. Nature 604, 502–508 (2022).

38. Petrovski, S., Wang, Q., Heinzen, E. L., Allen, A. S. & Goldstein, D. B. Genic Intolerance to Functional Variation and the Interpretation of Personal Genomes. PLOS Genet. 9, e1003709 (2013).

39. Lisabeth, E. M., Falivelli, G. & Pasquale, E. B. Eph Receptor Signaling and Ephrins. Cold Spring Harb. Perspect. Biol. 5, a009159 (2013).

40. Donoghue, M. J. & Rakic, P. Molecular gradients and compartments in the embryonic primate cerebral cortex. Cereb. Cortex N. Y. N 1991 9, 586–600 (1999).

41. Jayasena, C. S., Flood, W. D. & Koblar, S. A. High EphA3 expressing ophthalmic trigeminal sensory axons are sensitive to ephrin-A5-Fc: Implications for lobe specific axon guidance. Neuroscience 135, 97–109 (2005).

42. Wainberg, M. et al. Genetic architecture of the structural connectome. Nat. Commun. 15, 1962 (2024).

43. Di Palma, T. et al. Pax8 has a critical role in epithelial cell survival and proliferation. Cell Death Dis. 4, e729–e729 (2013).

44. Di Palma, T. et al. Characterization of a novel loss-of-function mutation of PAX8 associated with congenital hypothyroidism. Clin. Endocrinol. (Oxf.) 73, 808–814 (2010).

45. Mansouri, A., Chowdhury, K. & Gruss, P. Follicular cells of the thyroid gland require Pax8 gene function. Nat. Genet. 19, 87–90 (1998).

46. Son, S. M. et al. Thrombospondin-1 prevents amyloid beta-mediated synaptic pathology in Alzheimer’s disease. Neurobiol. Aging 36, 3214–3227 (2015).

47. Li, Z., Shue, F., Zhao, N., Shinohara, M. & Bu, G. APOE2: protective mechanism and therapeutic implications for Alzheimer’s disease. Mol. Neurodegener. 15, 63 (2020).

48. Kulminski, A. M., Philipp, I., Shu, L. & Culminskaya, I. Definitive roles of TOMM40-APOE-APOC1 variants in the Alzheimer’s risk. Neurobiol. Aging 110, 122–131 (2022).

49. Ken-Dror, G., Talmud, P. J., Humphries, S. E. & Drenos, F. APOE/C1/C4/C2 gene cluster genotypes, haplotypes and lipid levels in prospective coronary heart disease risk among UK healthy men. Mol. Med. Camb. Mass 16, 389–399 (2010).

50. ThrombospondinL1 binds to ApoER2 and VLDL receptor and functions in postnatal neuronal migration | The EMBO Journal. https://www.embopress.org/doi/full/10.1038/emboj.2008.223?pubCode=cgi.

51. Sensi, S. L., Paoletti, P., Bush, A. I. & Sekler, I. Zinc in the physiology and pathology of the CNS. Nat. Rev. Neurosci. 10, 780–791 (2009).

52. Chowanadisai, W., Graham, D. M., Keen, C. L., Rucker, R. B. & Messerli, M. A. Neurulation and neurite extension require the zinc transporter ZIP12 (slc39a12). Proc. Natl. Acad. Sci. 110, 9903–9908 (2013).

53. Scarr, E. et al. Increased cortical expression of the zinc transporter SLC39A12 suggests a breakdown in zinc cellular homeostasis as part of the pathophysiology of schizophrenia. Npj Schizophr. 2, 1–7 (2016).

54. Karczewski, K. J. et al. Systematic single-variant and gene-based association testing of thousands of phenotypes in 394,841 UK Biobank exomes. Cell Genomics 2, 100168 (2022).

55. Bullmore, E. & Sporns, O. The economy of brain network organization. Nat. Rev. Neurosci. 13, 336–349 (2012).

56. Foo, H. et al. Genetic influence on ageing-related changes in resting-state brain functional networks in healthy adults: A systematic review. Neurosci. Biobehav. Rev. 113, 98–110 (2020).

57. Bell, S., Tozer, D. J. & Markus, H. S. Genome-wide association study of the human brain functional connectome reveals strong vascular component underlying global network efficiency. Sci. Rep. 12, 14938 (2022).

58. van der Meer, D. et al. Boosting Schizophrenia Genetics by Utilizing Genetic Overlap With Brain Morphology. Biol. Psychiatry 92, 291–298 (2022).

59. Cao, H., Zhou, H. & Cannon, T. D. Functional connectome-wide associations of schizophrenia polygenic risk. Mol. Psychiatry 26, 2553–2561 (2021).

60. Carnevale, L. et al. Brain Functional Magnetic Resonance Imaging Highlights Altered Connections and Functional Networks in Patients With Hypertension. Hypertension 76, 1480–1490 (2020).

61. Hussein, A. et al. The association between resting-state functional magnetic resonance imaging and aortic pulse-wave velocity in healthy adults. Hum. Brain Mapp. 41, 2121– 2135 (2020).

62. Chang, C. et al. Association between heart rate variability and fluctuations in resting-state functional connectivity. NeuroImage 68, 93–104 (2013).

63. Wei, Y., et al. Connectome-wide Association of Polygenic Liability for Schizophrenia and Bipolar Disorder. Preprint at 10.31234/osf.io/z932j (2022).

64. Sjögren, M. et al. Tau and Aβ42 in Cerebrospinal Fluid from Healthy Adults 21–93 Years of Age: Establishment of Reference Values. Clin. Chem. 47, 1776–1781 (2001).

65. Slot, R. E. R. et al. Apolipoprotein A1 in Cerebrospinal Fluid and Plasma and Progression to Alzheimer’s Disease in Non-Demented Elderly. J. Alzheimers Dis. JAD 56, 687–697 (2017).

66. Yu, M., Sporns, O. & Saykin, A. J. The human connectome in Alzheimer disease — relationship to biomarkers and genetics. Nat. Rev. Neurol. 17, 545–563 (2021).

67. Jansen, P. R. et al. Genome-wide meta-analysis of brain volume identifies genomic loci and genes shared with intelligence. Nat. Commun. 11, 5606 (2020).

68. Nielsen, R., Paul, J. S., Albrechtsen, A. & Song, Y. S. Genotype and SNP calling from next-generation sequencing data. Nat. Rev. Genet. 12, 443–451 (2011).

69. Smith, S., Alfaro-Almagro, F. & Miller, K. L. UK Biobank Brain Imaging Documentation Version 1.9. (2022).

70. de Lange, S. C., Helwegen, K. & van den Heuvel, M. P. Structural and functional connectivity reconstruction with CATO - A Connectivity Analysis TOolbox. NeuroImage 273, 120108 (2023).

71. Jenkinson, M., Beckmann, C. F., Behrens, T. E. J., Woolrich, M. W. & Smith, S. M. FSL. NeuroImage 62, 782–790 (2012).

72. Desikan, R. S. et al. An automated labeling system for subdividing the human cerebral cortex on MRI scans into gyral based regions of interest. NeuroImage 31, 968–980 (2006).

73. Fischl, B. et al. Whole brain segmentation: automated labeling of neuroanatomical structures in the human brain. Neuron 33, 341–355 (2002).

74. Power, J. D., Barnes, K. A., Snyder, A. Z., Schlaggar, B. L. & Petersen, S. E. Spurious but systematic correlations in functional connectivity MRI networks arise from subject motion. NeuroImage 59, 2142–2154 (2012).

75. Alfaro-Almagro, F. et al. Confound modelling in UK Biobank brain imaging. NeuroImage 224, 117002 (2021).

76. Purcell, S. et al. PLINK: a tool set for whole-genome association and population-based linkage analyses. Am. J. Hum. Genet. 81, 559–575 (2007).

77. Abraham, G., Qiu, Y. & Inouye, M. FlashPCA2: principal component analysis of Biobank-scale genotype datasets. Bioinformatics 33, 2776–2778 (2017).

78. König, I. R., Loley, C., Erdmann, J. & Ziegler, A. How to Include Chromosome X in Your Genome-Wide Association Study. Genet. Epidemiol. 38, 97–103 (2014).

79. Fairley, S., Lowy-Gallego, E., Perry, E. & Flicek, P. The International Genome Sample Resource (IGSR) collection of open human genomic variation resources. Nucleic Acids Res. 48, D941–D947 (2020).

80. Giambartolomei, C. et al. Bayesian Test for Colocalisation between Pairs of Genetic Association Studies Using Summary Statistics. PLOS Genet. 10, e1004383 (2014).

81. Benner, C. et al. Prospects of Fine-Mapping Trait-Associated Genomic Regions by Using Summary Statistics from Genome-wide Association Studies. Am. J. Hum. Genet. 101, 539–551 (2017).

82. Benner, C. et al. FINEMAP: efficient variable selection using summary data from genome-wide association studies. Bioinformatics 32, 1493–1501 (2016).

83. Weeks, E. M. et al. Leveraging polygenic enrichments of gene features to predict genes underlying complex traits and diseases. Nat. Genet. 55, 1267–1276 (2023).

84. Liberzon, A. et al. Molecular signatures database (MSigDB) 3.0. Bioinforma. Oxf. Engl. 27, 1739–1740 (2011).

85. MacArthur, J. et al. The new NHGRI-EBI Catalog of published genome-wide association studies (GWAS Catalog). Nucleic Acids Res. 45, D896–D901 (2017).

