## Supplementary material for "The genetic landscape of heterogeneity in human functional brain connectivity": Supplmentary Notes and Figures

References of figures and tables for each supplementary note are labelled Supplementary Note (SN) Figure/Table. Supplementary Figures for the main text can be found at the end of this document. Supplementary Tables can be found in the accompanying excel file.

**Authors**

Bernardo de APC Maciel^1^, Marijn Schipper^1^, Cato Romero^2^, Christiaan de Leeuw^1^, Koen Helwegen^1^, Danielle Posthuma^1,3^, Jeanne E. Savage^1^, Martijn P. van den Heuvel^1,2^

^1^ Department of Complex Trait Genetics, Center for Neurogenomics and Cognitive Research, Amsterdam Neuroscience, Vrije Universiteit Amsterdam, Amsterdam, the Netherlands

^2^ Department of Child and Adolescent Psychiatry and Psychology, Section Complex Trait Genetics, Amsterdam Neuroscience, Vrije Universiteit Medical Center, Amsterdam UMC, Amsterdam, the Netherlands

^3^ Department of Clinical Genetics, Section Complex Trait Genetics, Amsterdam Neuroscience, Vrije Universiteit Medical Center, Amsterdam University Medical Centre, Amsterdam 1081 HZ, the Netherlands

**Table of Contents**

Supplementary Methods

Supplementary Note 1 - Sample Demographics

Supplementary Note 2 - Subnetwork Enrichment Analyses

Supplementary Note 3 - Validation of overlapping loci

Supplementary Note 4 - Disorder Effect Extent

Supplementary Note 5 - Reliability Analysis

References

### Supplementary Methods

#### Subject Exclusions & Quality Control

Data were derived from the UK Biobank (UKB), a population-based cohort sample of approximately 500,000 adults in the UK. Participants were invited for an initial in-person visit to a study assessment centre in 2006-2010, during which numerous physical measurements and surveys were administered and blood samples were collected for genotyping. Participant data was linked to medical records via the National Health Services, and a subset of participants also completed MRI scans and additional follow-up data collection in subsequent years. All participants provided written informed consent, those who revoked their consent at the time of the main analysis (January 2023) were removed from subsequent students. The UKB received ethical approval from the National Research Ethics Service Committee North West-Haydock (reference 11/NW/0382), and all study procedures were in accordance with the World Medical Association for medical research. Access to the UK Biobank data was obtained under application number 16406.

In the full UKB sample, 481 subjects with sex aneuploidy, 370 with discordant reported and chromosomal sex, and 195 with high degrees of relatedness were excluded from further analyses. Next, on the subset of subjects with one imaging visit available (n = 36,969), related subjects were excluded. Subjects with high levels of kinship (KING coefficient>0.4) and the most inferred relatives were removed until no two subjects were reported to be third-degree relatives (or closer), accounting for a total of 5,241 exclusions. Population stratification was controlled by correcting for principal genomic components calculated with FLASHPCA2.^1^ Principal components from the 1000 Genomes reference populations^2^ were projected onto the called genotypes available in the UKB data and all 2,034 subjects for whom the projected scores were the furthest to the average score of Europeans (i.e. Mahalanobis distance to the average of Europeans < 6$\sigma$) were excluded from further analysis. Finally, 1,341 subjects considered to be imaging outliers were excluded from the analysis. These were subjects for which average head motion parameters, signal-to-noise ratio or discrepancy between T1-weighted and fMRI scan deviated from the median more than five times their median absolute variation.^3^ These genotyping data were collected with two different array types - UK Biobank Axiom (UKBA) and the UK BiLEVE (UKBB) arrays - which cover 812,428 unique genetic markers and overlap 95% in SNP content.

#### Subnetwork Definition

The definition of resting-state networks (RSNs) based on the Yeo-Krienen 7-network atlas was carried out as described before.^4^ Briefly, the Yeo-Krienen atlas contains a parcellation map of 7 large-scale functional resting-state networks, including the visual, somatomotor, dorsal attention, ventral attention, limbic, frontoparietal, and default mode network. An annotation file of the 7 functional networks was included for the fsaverage subject in the FreeSurfer Software package. For this, the surface-based annotation was translated to a 3D brain volume in volumetric space, in which each grey matter voxel was assigned a network label. Next, for each region in the Desikan-Killiany atlas, the ratio of voxels that belonged to each of the 7 RSNs was computed.^5^ Using a majority vote approach, the label of the functional network corresponding to the majority of voxels was then assigned to that region.

All edges connecting areas belonging to the same network were assigned to it. The following regions in the aseg atlas were assigned to the subcortical network (all bilateral): Thalamus, Caudate, Putamen, Pallidum, Hippocampus, Amygdala, Accumbens-area.^6^ Edges connecting cortical and subcortical edges were assigned to the cortical-subcortical category and the remaining to the between-network category. In total, the edges were divided into 10 different disjoint sets. The comprehensive list of subnetwork-edge pairings can be found in **Sup. Tab. 1**.

#### Subnetwork Enrichment Analysis

We performed enrichment analysis to identify whether any particular subset of edges in a given RSN presents a statistically significant difference in SNP-heritability. The heritability estimation for each edge was divided by its standard deviation to create a Z-score. These scores were not independent of each other because (i) edges in the same RSN are more similar than those outside of the RSN; (ii) the data has strong spatial dependencies. Therefore, not accounting for these spatial dependencies (e.g. two-sample t-tests or simple linear regressions) would lead to a highly inflated type I error rate.

For each RSN, we used a linear regression $Z = \beta_{0} + \beta_{s}X_{s} + \beta_{r}X_{r} + \epsilon_{r}$ on the$n = 3321$ edges as data points, with $Z$ the vector of Z-scores for each edge, $X_{s}$ the average edge strength, and $X_{r}$ a binary indicator variable scored 1 if the edge belonged to that RSN and 0 otherwise. The residuals were modelled as$\epsilon_{r} \sim MVN(0, {\sigma_{r}}^{2}S)$, with ${\sigma_{r}}^{2}$ the residual variance parameter. The fixed $n$ x $n$ correlation matrix $S$ was included to account for the dependencies between the heritability estimates of the edges, and was set to the element-wise square of the phenotypic correlation matrix of the edges in the discovery sample.

The models were fitted using Generalized Least Squares. The parameter for each RSN represents the mean difference in strength of genetic signal (as quantified by their Z-score) of edges in that RSN compared to the other edges, after correcting for average edge strength, with positive corresponding to an enrichment of genetic signal in the RSN. Consequently, we performed a one-sided test of $H_{0}: \beta_{r}=0$ against the alternative hypothesis $H_{1}: \beta_{r}>0$, using a t-test with 269 degrees of freedom (derived from the rank of $S$).

### Supplementary Note 1 - Sample Demographics

After quality control, a total of 28,159 subjects of European ancestry were considered for this study. These comprised a discovery and replication sample of 24,451 and 3,708 unique unrelated individuals, respectively. A complete description of the sample demographic characteristics can be found in **Supplementary Note (SN) Table S1**. Field codes for all variables used in this study can be found on **SN Tab. S2**. Age, head motion and intracranial volume did not follow a normal distribution (**SN Fig. S1**). Association testing showed a small significant difference between groups for age. The standardised mean difference for age between groups was 0.182, which was considered a small effect size and unlikely to confound the genetic analyses.

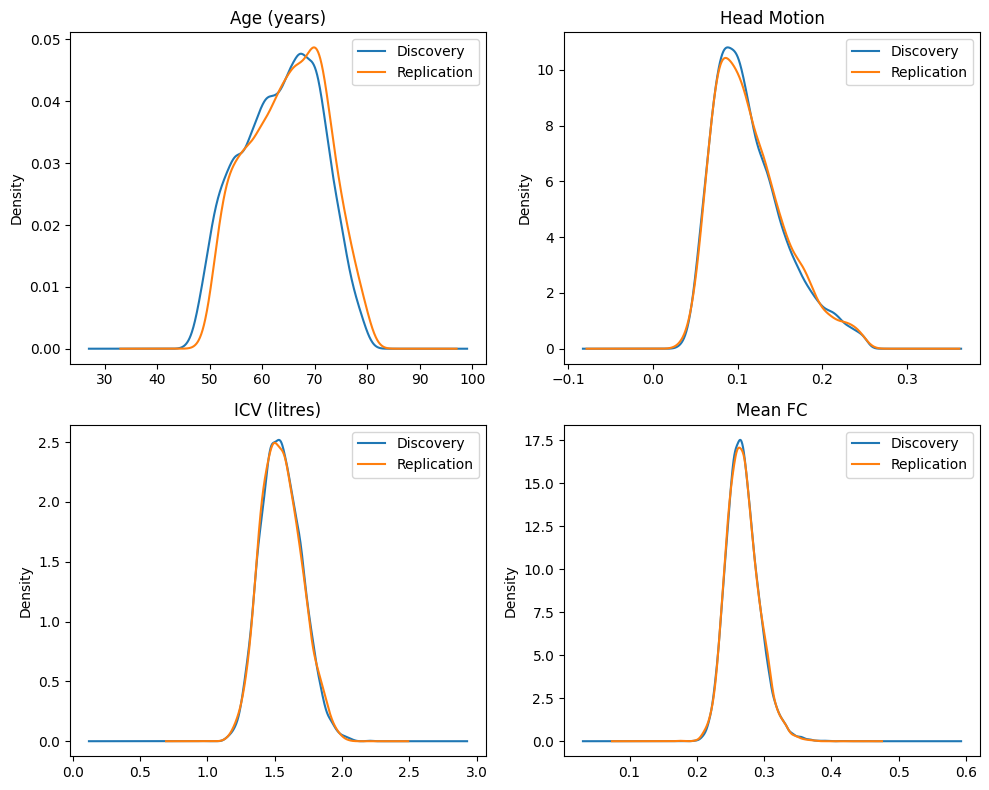

**Supplementary Note Figure S1. Density plots for non-normal demographic variables.** Age (top left), Head Motion (top right), Mean of Non-Negative functional connections (bottom left) and Intracranial Volume (bottom right). The density curves are plotted in blue for the discovery and orange for the replication samples.

**Supplementary Note Table 1. Description of the demographic sample and significance of association testing.** Chromosomal sex, handedness and array type are categorical variables, association was tested using the chi-squared test. The Kruskal-Wallis H test calculates p-values for age, head motion and intracranial volume sample differences. Reported p-values are Bonferroni-corrected for the number of tests. Q1: first quartile; Q3: third quartile; UKBA: UK Biobank Axiom; UKBB: UK BiLEVE.

|  |  | **Overall** | **Discovery** | **Replication** | **P-Value (adjusted)** |
| --- | --- | --- | --- | --- | --- |
| **N** |  | 28159 | 24451 | 3708 |  |
| **Chromosomal Sex, N (%)** | Female | 15197 (54.0) | 13151 (53.8) | 2046 (55.2) | 0.818 |
|  | Male | 12962 (46.0) | 11300 (46.2) | 1662 (44.8) |  |
| **Age, median [Q1,Q3]** |  | 64.0 [58.0,70.0] | 64.0 [58.0,69.0] | 65.0 [59.0,71.0] | <0.001 |
| **Head Motion, median [Q1,Q3]** |  | 0.1 [0.1,0.1] | 0.1 [0.1,0.1] | 0.1 [0.1,0.1] | 1.000 |
| **Handedness, N (%)** | Left | 2653 (9.4) | 2269 (9.3) | 384 (10.4) | 0.276 |
|  | Non-left | 25506 (90.6) | 22182 (90.7) | 3324 (89.6) |  |
| **Array Type, N (%)** | UKBA | 25653 (91.1) | 22273 (91.1) | 3380 (91.2) | 1.000 |
|  | UKBB | 2506 (8.9) | 2178 (8.9) | 328 (8.8) |  |
| **Intracranial Volume (litres), median [Q1,Q3]** |  | 1.5 [1.4,1.7] | 1.5 [1.4,1.7] | 1.5 [1.4,1.7] | 1.000 |
| **Mean Functional Connectivity, median [Q1,Q3]** |  | 0.3 [0.3,0.3] | 0.3 [0.3,0.3] | 0.3 [0.3,0.3] | 1.000 |

**Supplementary Note Table 2. UKB field codes for covariates.**

| **Field** | **Description** |
| --- | --- |
| f.31.0.0 | Self-reported sex |
| f.54.2.0 | Screening and Imaging Centre |
| f.21003.2.0 | Age at imaging visit |
| f.1707.0.0 | Handedness |
| f.22000.0.0 | Genomic batch |
| f.25744.2.0 | Signal-to-Noise Ratio |
| f.25756.2.0 | Position of the table in the X axis |
| f.25757.2.0 | Position of the table in the Y axis |
| f.25758.2.0 | Position of the table in the Z axis |
| f.25759.2.0 | Position of the coil relative to table |
| f.25923.2.0 | Time-to-Echo |
| f.25929.2.0 | Intensity Scaling |
| f.26521.2.0 | Intracranial Volume |

### Supplementary Note 2 - Subnetwork Enrichment Analyses

In addition to the analyses on subnetwork enrichment (**Results**; **Sup. Tab. 1**), enrichment in intrahemispheric, interhemispheric, subcortical and cortico-subcortical edges (referred to as edge topology; **SN Tab. S3**) was also calculated. None of the enrichment values was significant. **Supplementary Note Figure 2** plots the distribution of the heritability estimates grouped by topology.

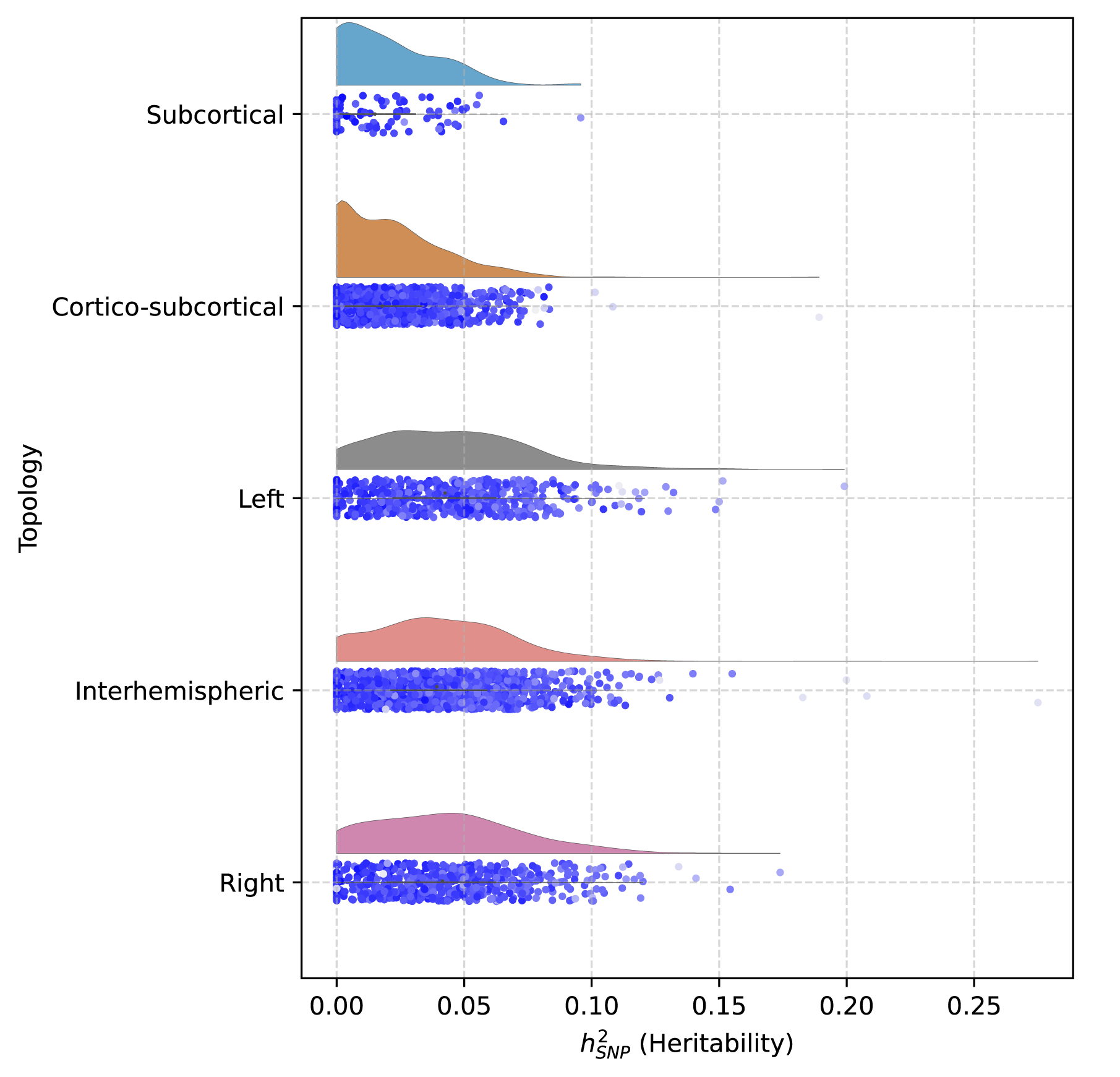

**Supplementary Note Figure 2. Raincloud plots for heritability estimates grouped per topology.** Half-violin plots represent the distribution of edge LDSC SNP-heritability. Each point below the violin plot is the heritability estimate for each edge belonging to the network. The shade of points in the jitter plot is the standard deviation of the estimation: lighter - higher uncertainty.

**Supplementary Note Table 3. Enrichment for different topologies of edges.** ${h^{2}}_{SNP}$ represents the average SNP-heritability estimate of the edges in the subnetwork (mean ± standard error). $N_{Edges}$ is the number of edges in each subnetwork.

| Topology | ${h^{2}}_{SNP}$ | $N_{Edges}$ | Enrichment  $t(269)$ | P-value |
| --- | --- | --- | --- | --- |
| Interhemispheric | 0.042 ± 0.029 | 1156 | 0.497 | .310 |
| Left | 0.044 ± 0.029 | 561 | 0.275 | .392 |
| Right | 0.043 ± 0.030 | 561 | -0.675 | .641 |
| Subcortical | 0.019 ± 0.019 | 91 | -0.820 | .794 |

### Supplementary Note 3 - Validation of overlapping loci

SNP prioritisation and loci co-localisation were carried out to ensure the overlapping loci across GWAS have the same source of genetic signal. The determination of common likely causal SNPs for different locus-edge associations was performed by colocalising overlapping loci. For this purpose, two loci from two different GWAS were considered to overlap if their respective lead SNPs were separated by less than 250K base pairs. If a locus was small (<250K base pairs) a 250K region around the center of the locus was considered (**SN Fig. 3**). A total of 79 replicated locus-edge were found to be associated with three overlapping loci: 11 with *PAX8* (2:113963070:114213070), 13 with *EphA3* (3:89451721:90010903) and 3 with *THBS1* (15:39514832:39764832). Summary statistics in the overlapping loci were filtered to contain only SNPs that reach the marginal significance threshold of $\alpha\boldsymbol{=0.05}$ in the three loci of interest. Estimation of the LD reference panel consisting of all the individuals in each discovery GWAS was carried out on LDStore2 (v2.0).^7^ The resulting LD reference panel was used to fine-map each of the overlapping loci with FINEMAP (v1.4.1), using a threshold of a maximum of 10 causal variants per locus.^8^ Furthermore, the probability of the overlapping loci having the same causal variant across GWAS was estimated using approximate Bayes factor co-localisation, implemented in coloc (v5.2.2).^9^ Two locus-trait associations were considered colocalised if these had a 90% probability of sharing a single causal variant ($\boldsymbol{H}_{\boldsymbol{4}}\boldsymbol{>0.9}$).

For the three overlapping loci, all 78 locus-trait associations were predicted to have only one causal variant. For the *PAX8* and *THBS1* loci, all edges were found to be co-localised ($\boldsymbol{H}_{\boldsymbol{4}}\boldsymbol{>0.97}$ for all comparisons). For the EphA3 locus, 85% of the 11 edges co-localised (66 out of 78 unique edge pairs). The associations that did not co-localise were always with the intrahemispheric connectivity between the right supramarginal and medial temporal gyri (**SN Fig. 4**). All of these had moderate (> 45%) $\boldsymbol{H}_{\boldsymbol{4}}$ probabilities. In case $\boldsymbol{H}_{\boldsymbol{4}}\boldsymbol{< 0.9}$, $\boldsymbol{H}_{\boldsymbol{3}}$would be the second highest probability by several orders of magnitude. These all happened for the same edge, indicating that this edge is likely associated with a different SNP in this locus (**SN Fig. 5**). However, FLAMES mapped the same gene to this locus, showing that downstreams effects are likely to be converging on the same gene. Taken together, the results reflected a high probability that the genetic effects we observed in the 22 individual edge-GWAS were all associated with the same genetic variants within the three overlapping loci. This brings further evidence for the pleiotropy of these discovered loci for several functional links across the human brain.

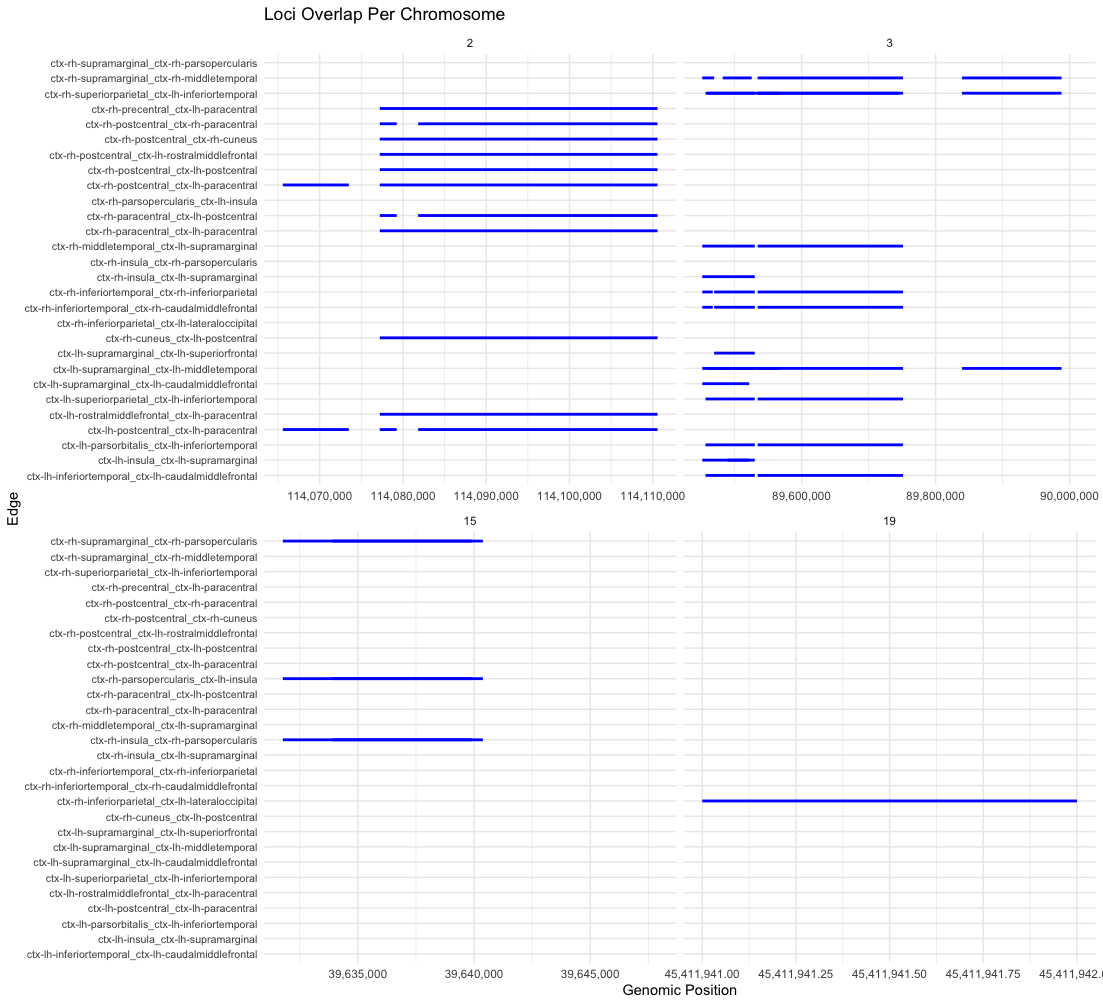

**Supplementary Note Figure 3. Visualisation of locus boundary overlap.** Each blue line represents one significant replicated locus association with the respective edge. Locus edge associations are found in close proximity across the four loci, being mapped into one overlapping locus encompassing the different locus-edge associations.

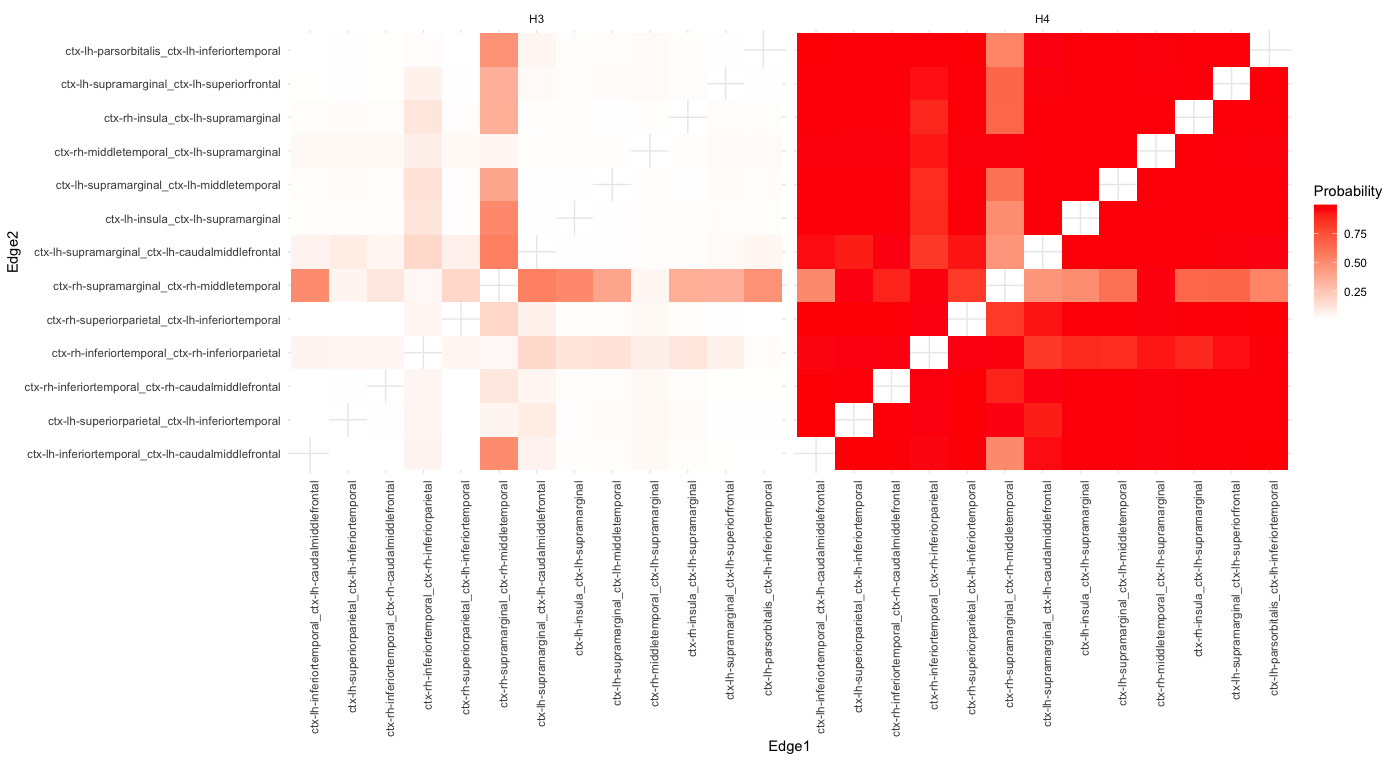

**Supplementary Note Figure 4. Heatmap for** $\boldsymbol{H}_{\boldsymbol{3}}$ **(left) and** $\boldsymbol{H}_{\boldsymbol{4}}$ **(right) probability in coloc for all unique pairs of edges associated with the EphA3 locus (3:89451721:90010903).** All pairs of edges are found to have a high probability of having the same putative causal variant, except for the right supramarginal and medial temporal gyri (right). This edge is found to have a modest probability of having a unique causal variant in this locus (left).

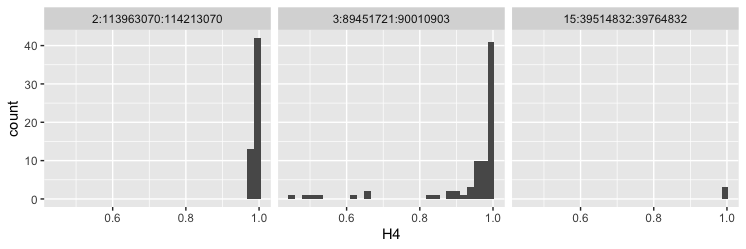

**Supplementary Note Figure 5. Histograms for** $\boldsymbol{H}_{\boldsymbol{4}}$ **probability in coloc for all unique pairs of edges associated with the *PAX8* (2:113963070:114213070), *EphA3* (3:89451721:90010903) and *THBS1* locus (15:39514832:39764832).** Locus positions according to GRCh37 build.

### Supplementary Note 4 - Disorder Effect Extent

#### Gene-sets for disorder

All most recent case-control disorder GWAS per working group in the Psychiatric Genetic Consortium were considered for this study. Summary statistics were uploaded to FUMA, where MAGMA was used to run GWGAS for all protein-encoding genes with a window of 0Kb around the gene and using the 1KG EUR reference panel as an LD reference.^2,10,11^ GWGAS with less than 5 significant genes were not kept for downstream analyses because of the small size of the gene-set. After this filtering, total of 8 different datasets were kept: Attention Deficit/Hyperactivity Disorder (ADHD; **SN Fig. 5**),^12^ Anorexia (**SN Fig. 6**),^13^ Autism Spectrum Disorder (ASD; **SN Fig. 7**),^14^ Alzheimer’s Disorder (AD; **SN Fig. 8**),^15^ Bipolar Disorder (BIP; **SN Fig. 9**),^16^ Depression (Dep; **SN Fig. 10**),^17^ Substance Use Disorder (SUD; **SN Fig. 11**)^18^ and Schizophrenia (**SN Fig. 12**).^19^ A gene-set was built for each disorder by joining all the genes with a p-value lower than 0.05/18623 (corrected for number of genes tested).

#### Gene effect extent

The effect extent for each gene was calculated using a procedure similar to that applied to loci (**Methods**). MAGMA was run for all edges of the connectome. SNP-wise mean model using an ancestry-specific reference panel derived from 10K unrelated individuals from the UK Biobank was applied to test the joint association of all SNPs within 18,850 protein-encoding genes with each trait. Results were aggregated per gene across all GWGAS and Benjamini-Hochberg FDR correction for the number of functional connectivity traits (3,321) was applied to these p-values to quantify with how many traits each gene was expected to be associated. This number is referred to as the effect extent of a gene. To calculate the average effect extent of a gene-set, the effect extent of all the genes included in this gene-set were averaged.

#### Permutation testing and null models

A gene-gene correlation (LD) matrix was obtained by running MAGMA on the functional connectivity phenotypes. Fixed columns of the .raw MAGMA file were discarded and the partial correlation matrix was transformed into a sparse matrix representing LD between genes ($\Sigma$).

Three null models were considered to compare the effect extent of disease genes against a background of random genes: LD-aware gene-sets of random genes, and LD-aware and LD-agnostic gene-sets of highly brain expressed genes. First, a simple LD-agnostic background of all highly expressed brain genes as defined in the background of SynGO (<https://www.syngoportal.org/data/background_brain.json>) by sampling N random genes, where N is the number of genes significant for a given phenotype.^20^ LD-aware aware models were defined based on the sampling of a multivariate normal distribution with a root matrix derived from $\Sigma$. Per phenotype, 18,623 (for model with all genes) or 6,401 (model with highly expressed brain genes) random values were sampled out of $Z\sim MVN(0,\Sigma)$. These values were ranked and the first N genes were picked. In this way, the random sampling of the values reflects the LD structure between genes. Each of these three sampling procedures was repeated 50,000 times per phenotype to generate null models for the average extent of the effect of a random gene-set on the functional connectome.

#### Disorder gene-set effect extent comparison

Permutation testing was performed to determine whether disorder genes are expected to affect more of the functional connectome than random genes. On average, if a gene is known to be involved in disorder, it was found to be affecting more of the functional connectome for most brain disorders: ADHD (p = .04; p-value for a random LD-aware gene-set of brain expressed genes), anorexia (p = .03), Alzheimer’s (p < .0001), BIP (p = .02) and Scz (p < .0001). After correcting for multiple testing for the number of disorders, the genesets for AD and SCZ were still significant for having a larger effect on the connectome than a LD-equivalent set of highly brain-expressed genes. Results are summarised in **Sup. Fig. 5** and **SN Table 4**. These results strengthen the relationship between genes, connectivity and disorder, showing that there is pervasive pleiotropy between neuropsychiatric disorders and the resting-state of the brain.

**Supplementary Note Figure 6. ADHD GWGAS.** Significant genes are annotated. Horizontal line denotes significance level 0.05/18623 = 2.651 x 10^-6^.

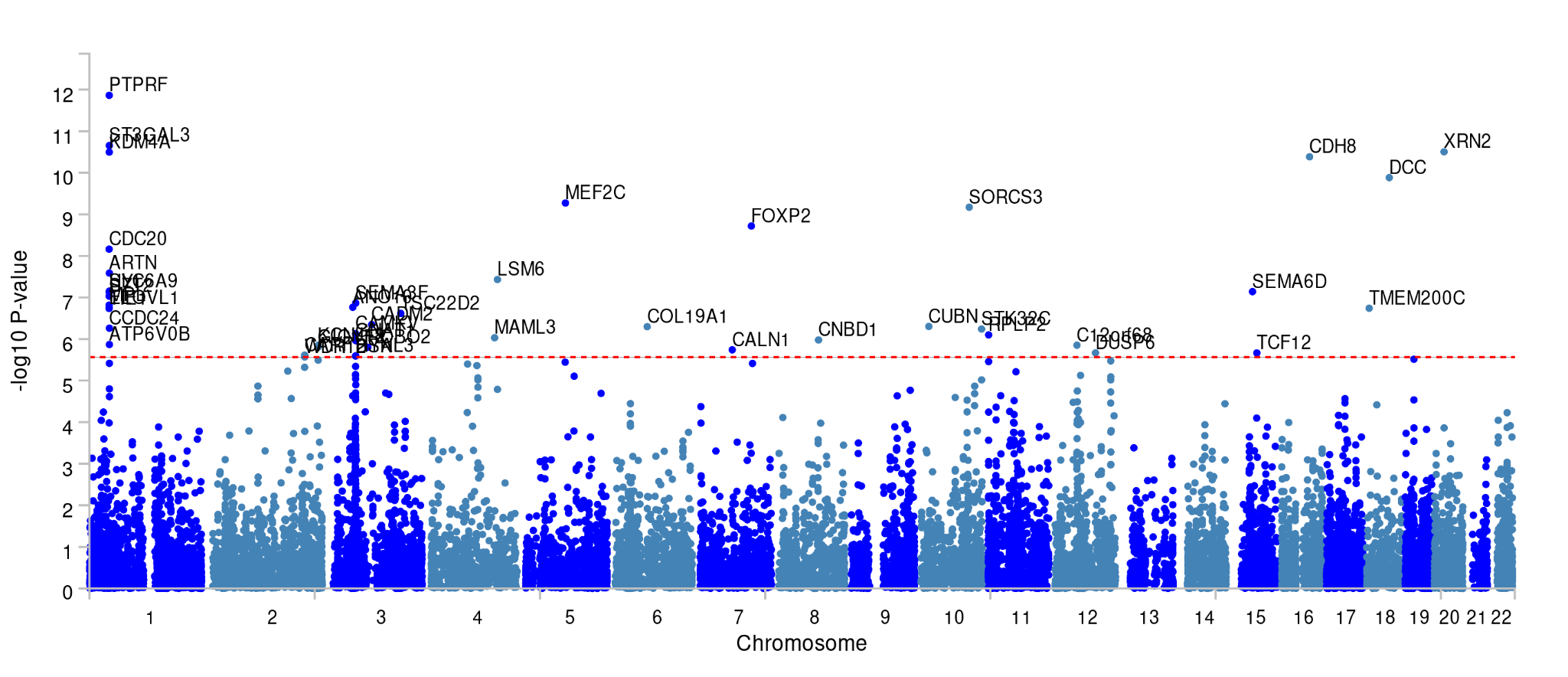

**Supplementary Note Figure 7. Anorexia GWGAS.** Significant genes are annotated. Horizontal line denotes significance level 0.05/18623 = 2.651 x 10^-6^.

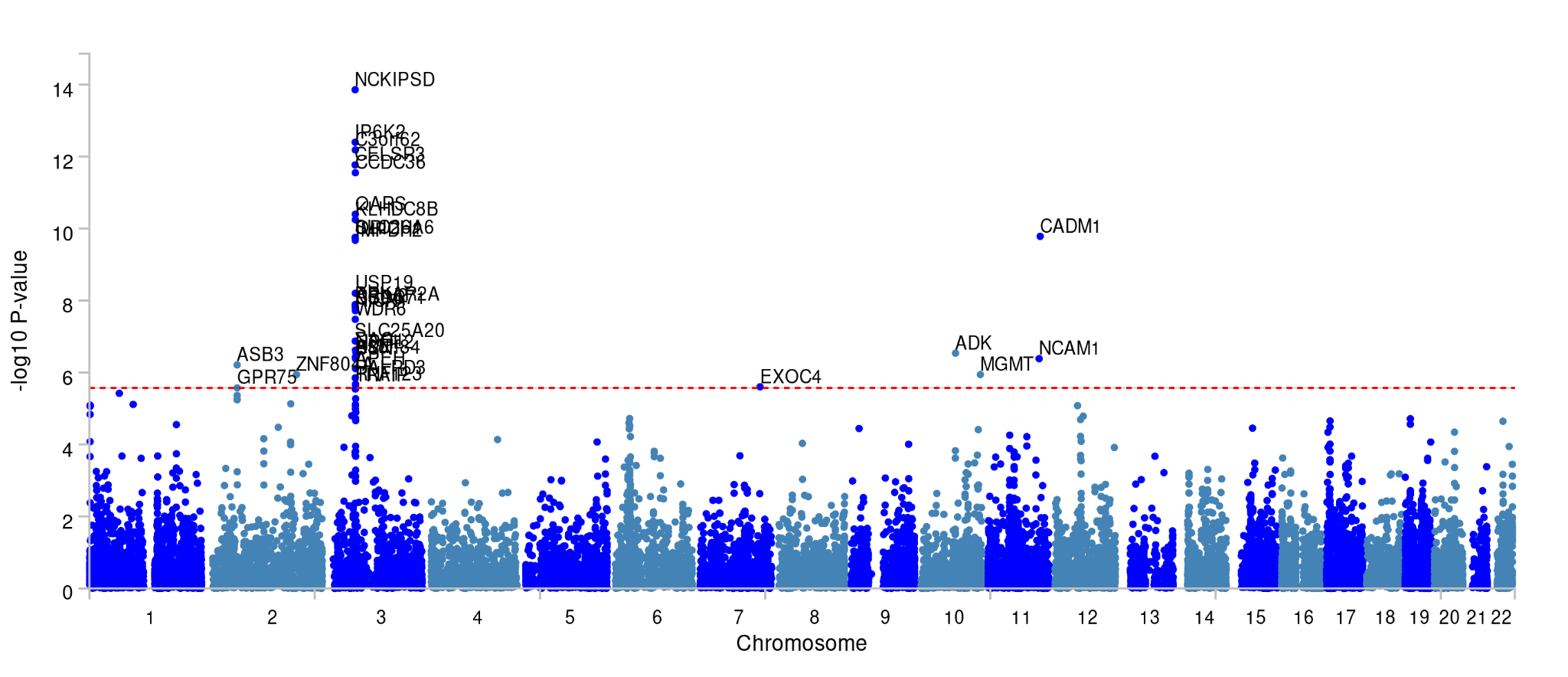

**Supplementary Note Figure 8. Alzheimer’s Disorder GWGAS.** Significant genes are annotated. Horizontal line denotes significance level 0.05/18623 = 2.651 x 10^-6^.

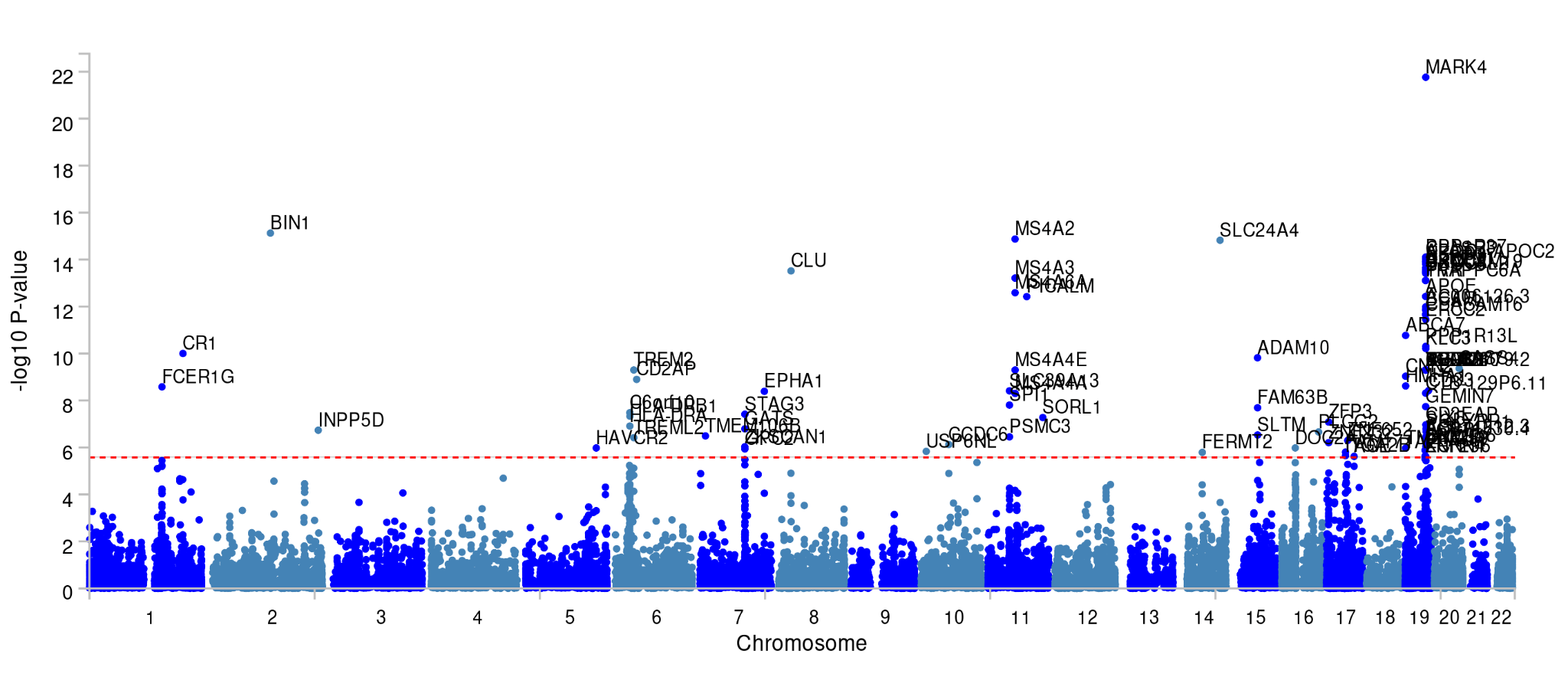

**Supplementary Note Figure 9. Bipolar Disorder GWGAS.** Significant genes are annotated. Horizontal line denotes significance level 0.05/18623 = 2.651 x 10^-6^.

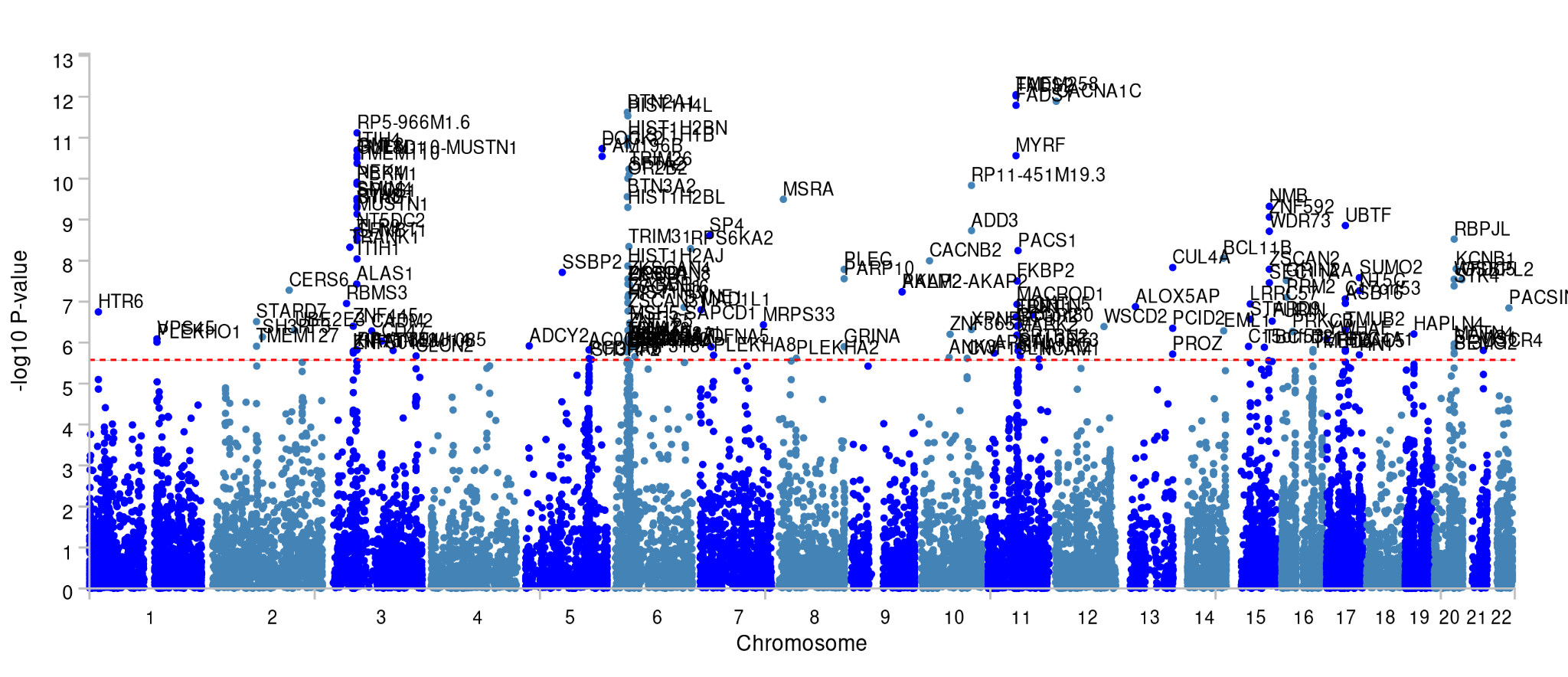

**Supplementary Note Figure 10. Depression GWGAS.** Significant genes are annotated. Horizontal line denotes significance level 0.05/18623 = 2.651 x 10^-6^.

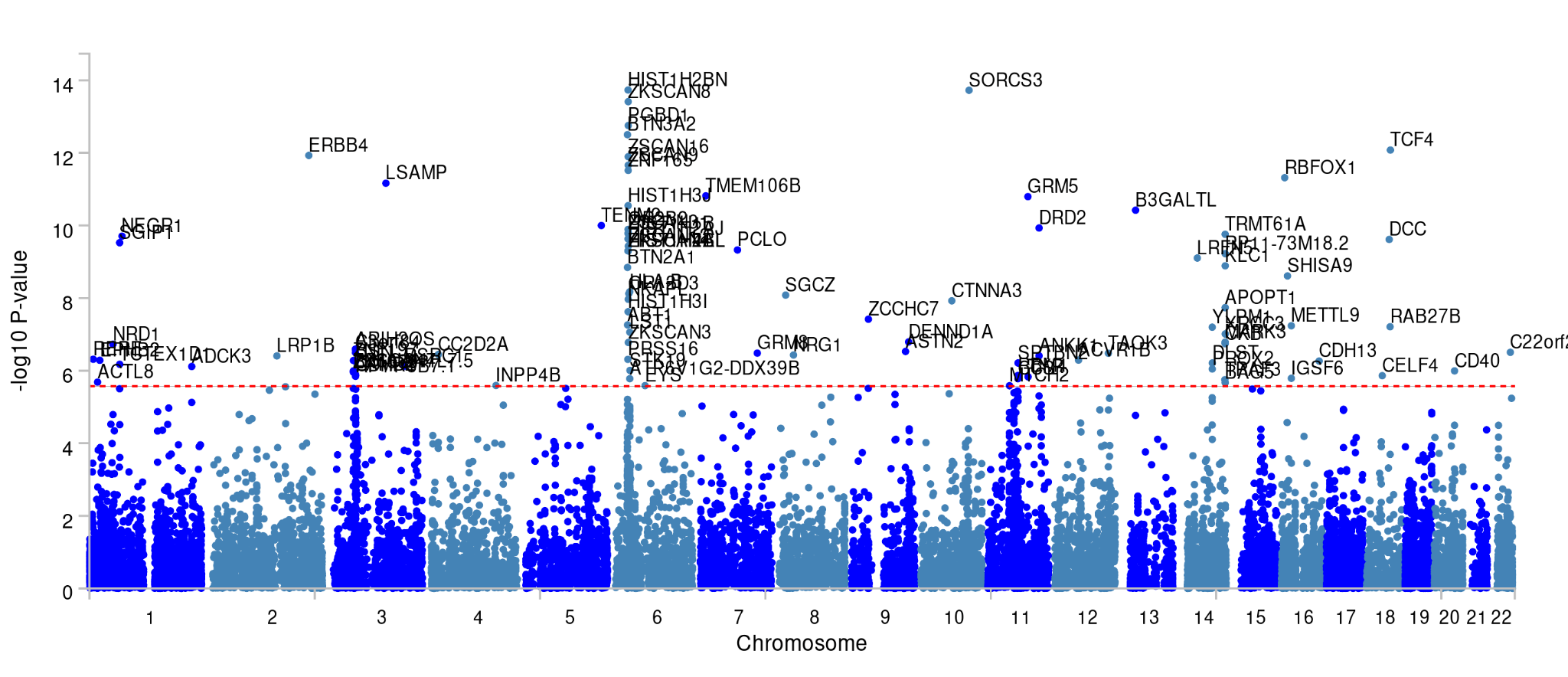

**Supplementary Note Figure 11. Substance Use Disorder GWGAS.** Significant genes are annotated. Horizontal line denotes significance level 0.05/18623 = 2.651 x 10^-6^.

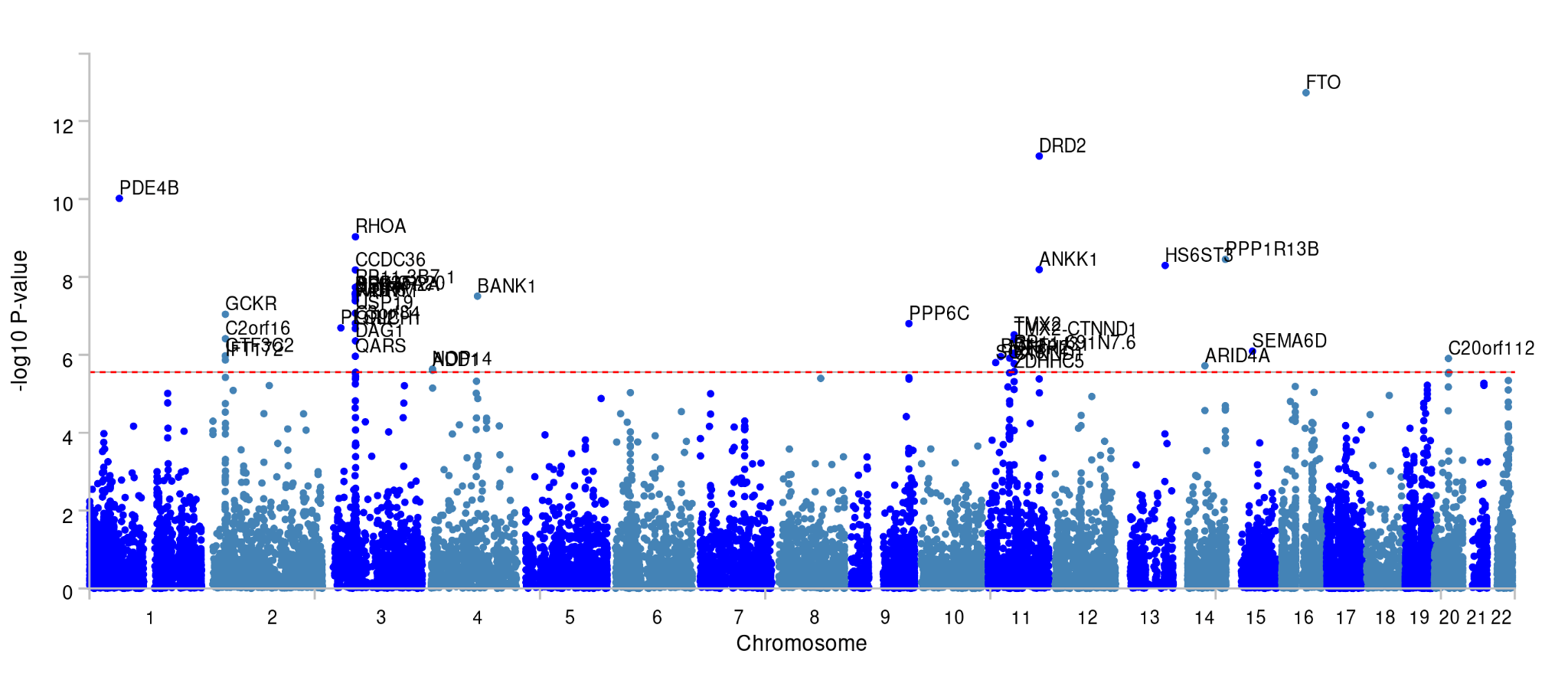

**Supplementary Note Figure 12. Schizophrenia GWGAS.** Top 100 Significant genes are annotated. Horizontal line denotes significance level 0.05/18623 = 2.651 x 10^-6^.

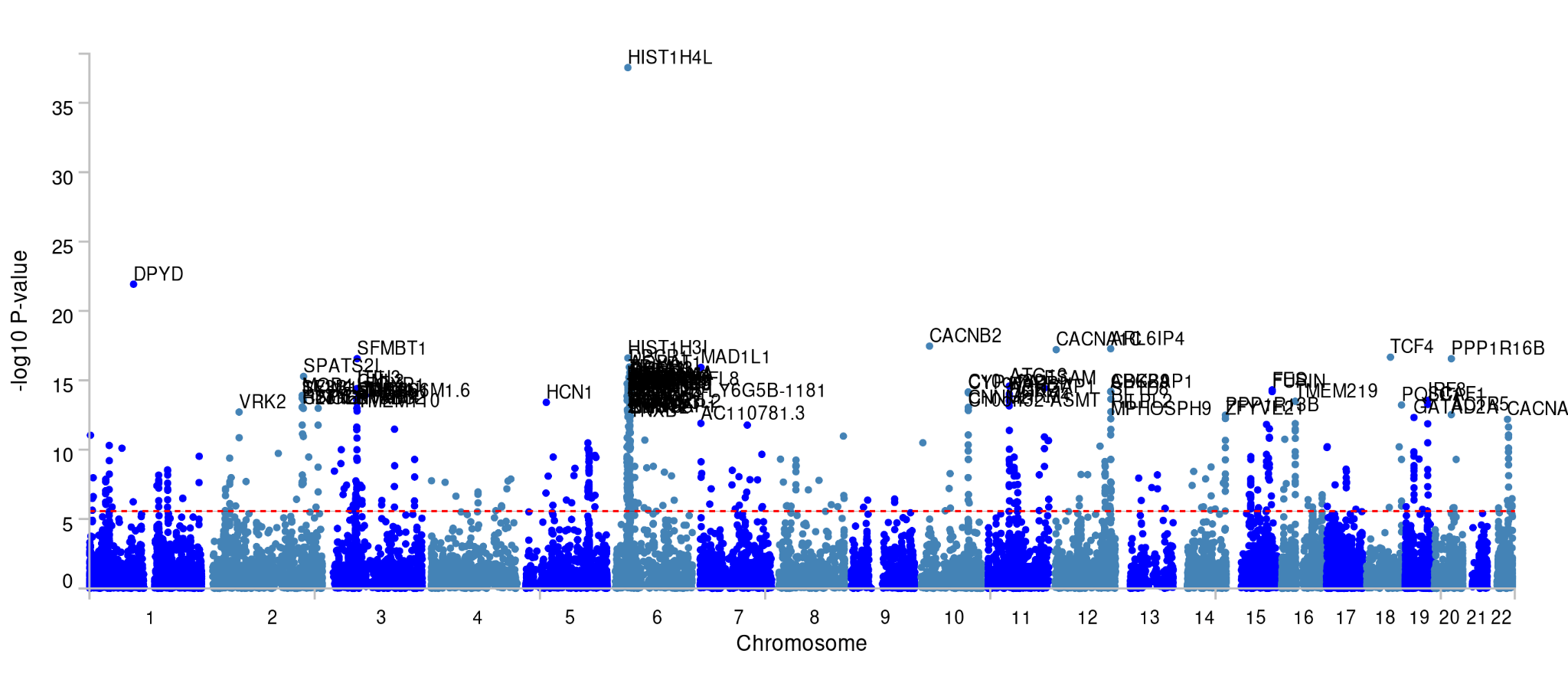

**Supplementary Note Table 4. Permutation testing results for the effect extent of different neuropsychiatric disorders.** P-values for the different null models: LD-aware with a background of all protein-encoding genes and LD-agnostic and LD-aware with a background of highly brain expressed genes. $N_{genes}$ is the number of significant genes per disorder.

| Source GWAS | $N_{Genes}$ | $p$-value LD-agnostic brain | $p$-value LD-aware protein-encoding | $p$-value LD-aware brain |
| --- | --- | --- | --- | --- |
| ADHD (Demontis 2022) | 46 | .021 | .026 | .036 |
| Anorexia (Watson 2019) | 38 | .013 | .018 | .028 |
| AD (Wightman 2021) | 97 | >.0001 | >.0001 | >.0001 |
| BIP (Mullins 2021) | 216 | .056 | .065 | .093 |
| Depression (Howard 2019) | 133 | .057 | .060 | .091 |
| SUD (Hatoum 2023) | 42 | .287 | .206 | .267 |
| Scz (Trubetskoy 2022) | 753 | >.0001 | >.0001 | >.0001 |

### Supplementary Note 5 - Reliability Analysis

The relationship between the reliability of edges and heritability was investigated to characterise the power in this study. From our total sample of 28,159 individuals, 3,018 had a second scan available with similar preprocessing. The intraclass correlation coefficient ICC(3,1) was calculated for each of the 3,321 edges and used to access univariate test-retest reliability.

The ICC values ranged from -0.05 to 0.75 with a median of 0.32. Spearman’s correlation between the z-values of the LDSC SNP-heritability and ICC values were calculated to be 0.68, indicating a high correlation between the certainty of the SNP-heritability estimation and the reliability of the phenotypic measurement (**SN Fig. 13**). These results showed heritability estimates are higher for those edges with the highest measurement reliability, meaning that noisy measurements tend to yield lower heritability estimates reflecting a relative loss of power. Despite the substantial correlation between these two sets of values, measurement reliability is not the only factor influencing the estimation of SNP-heritability, as different edges of the connectome may be affected by genes and the environment to different extents.

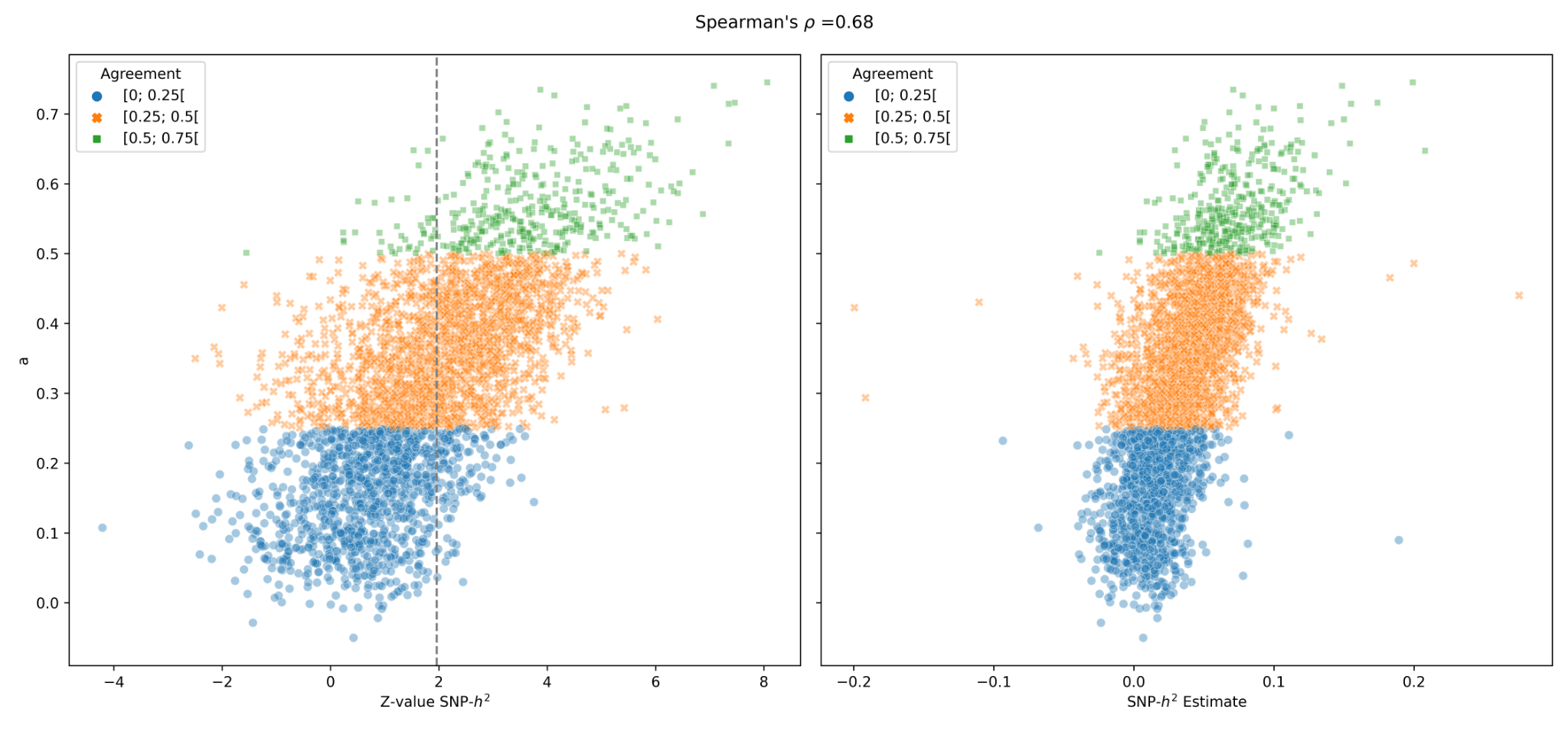

**Supplementary Note Figure 13. Relationship between measurement reliability and SNP-heritability.** Left| Comparison with Z-value of the SNP-h2 estimation. The grey dashed line represents the threshold for significant heritability. Measurements with higher reliability tend to have higher and more significant estimations of SNP-heritability. Right| Comparison with SNP-h2 estimation. Note that the heritability estimates are highly variable, reflecting a possibly different contribution of common variants to different brain connections, even among those for which the measurement is the most reliable (green class).

### References

1.	Abraham, G., Qiu, Y. & Inouye, M. FlashPCA2: principal component analysis of Biobank-scale genotype datasets. *Bioinformatics* **33**, 2776–2778 (2017).

2.	Fairley, S., Lowy-Gallego, E., Perry, E. & Flicek, P. The International Genome Sample Resource (IGSR) collection of open human genomic variation resources. *Nucleic Acids Res.* **48**, D941–D947 (2020).

3.	Alfaro-Almagro, F. *et al.* Confound modelling in UK Biobank brain imaging. *NeuroImage* **224**, 117002 (2021).

4.	Wei, Y. *et al.* Genetic mapping and evolutionary analysis of human-expanded cognitive networks. *Nat. Commun.* **10**, 4839 (2019).

5.	Desikan, R. S. *et al.* An automated labeling system for subdividing the human cerebral cortex on MRI scans into gyral based regions of interest. *NeuroImage* **31**, 968–980 (2006).

6.	Fischl, B. *et al.* Whole brain segmentation: automated labeling of neuroanatomical structures in the human brain. *Neuron* **33**, 341–355 (2002).

7.	Benner, C. *et al.* Prospects of Fine-Mapping Trait-Associated Genomic Regions by Using Summary Statistics from Genome-wide Association Studies. *Am. J. Hum. Genet.* **101**, 539–551 (2017).

8.	Benner, C. *et al.* FINEMAP: efficient variable selection using summary data from genome-wide association studies. *Bioinformatics* **32**, 1493–1501 (2016).

9.	Giambartolomei, C. *et al.* Bayesian Test for Colocalisation between Pairs of Genetic Association Studies Using Summary Statistics. *PLOS Genet.* **10**, e1004383 (2014).

10.	de Leeuw, C. A., Mooij, J. M., Heskes, T. & Posthuma, D. MAGMA: Generalized Gene-Set Analysis of GWAS Data. *PLoS Comput. Biol.* **11**, e1004219 (2015).

11.	Watanabe, K., Taskesen, E., van Bochoven, A. & Posthuma, D. Functional mapping and annotation of genetic associations with FUMA. *Nat. Commun.* **8**, 1826 (2017).

12.	Demontis, D. *et al.* Genome-wide analyses of ADHD identify 27 risk loci, refine the genetic architecture and implicate several cognitive domains. *Nat. Genet.* **55**, 198–208 (2023).

13.	Watson, H. J. *et al.* Genome-wide association study identifies eight risk loci and implicates metabo-psychiatric origins for anorexia nervosa. *Nat. Genet.* **51**, 1207–1214 (2019).

14.	Grove, J. *et al.* Identification of common genetic risk variants for autism spectrum disorder. *Nat. Genet.* **51**, 431–444 (2019).

15.	Wightman, D. P. *et al.* A genome-wide association study with 1,126,563 individuals identifies new risk loci for Alzheimer’s disease. *Nat. Genet.* **53**, 1276–1282 (2021).

16.	Mullins, N. *et al.* Genome-wide association study of more than 40,000 bipolar disorder cases provides new insights into the underlying biology. *Nat. Genet.* **53**, 817–829 (2021).

17.	Howard, D. M. *et al.* Genome-wide meta-analysis of depression identifies 102 independent variants and highlights the importance of the prefrontal brain regions. *Nat. Neurosci.* **22**, 343–352 (2019).

18.	Hatoum, A. S. *et al.* Multivariate genome-wide association meta-analysis of over 1 million subjects identifies loci underlying multiple substance use disorders. *Nat. Ment. Health* **1**, 210–223 (2023).

19.	Trubetskoy, V. *et al.* Mapping genomic loci implicates genes and synaptic biology in schizophrenia. *Nature* **604**, 502–508 (2022).

20.	Koopmans, F. *et al.* SynGO: An Evidence-Based, Expert-Curated Knowledge Base for the Synapse. *Neuron* **103**, 217-234.e4 (2019).

### Supplementary Figures

**Supplementary Figure 1. Visual summary of methods and results.**

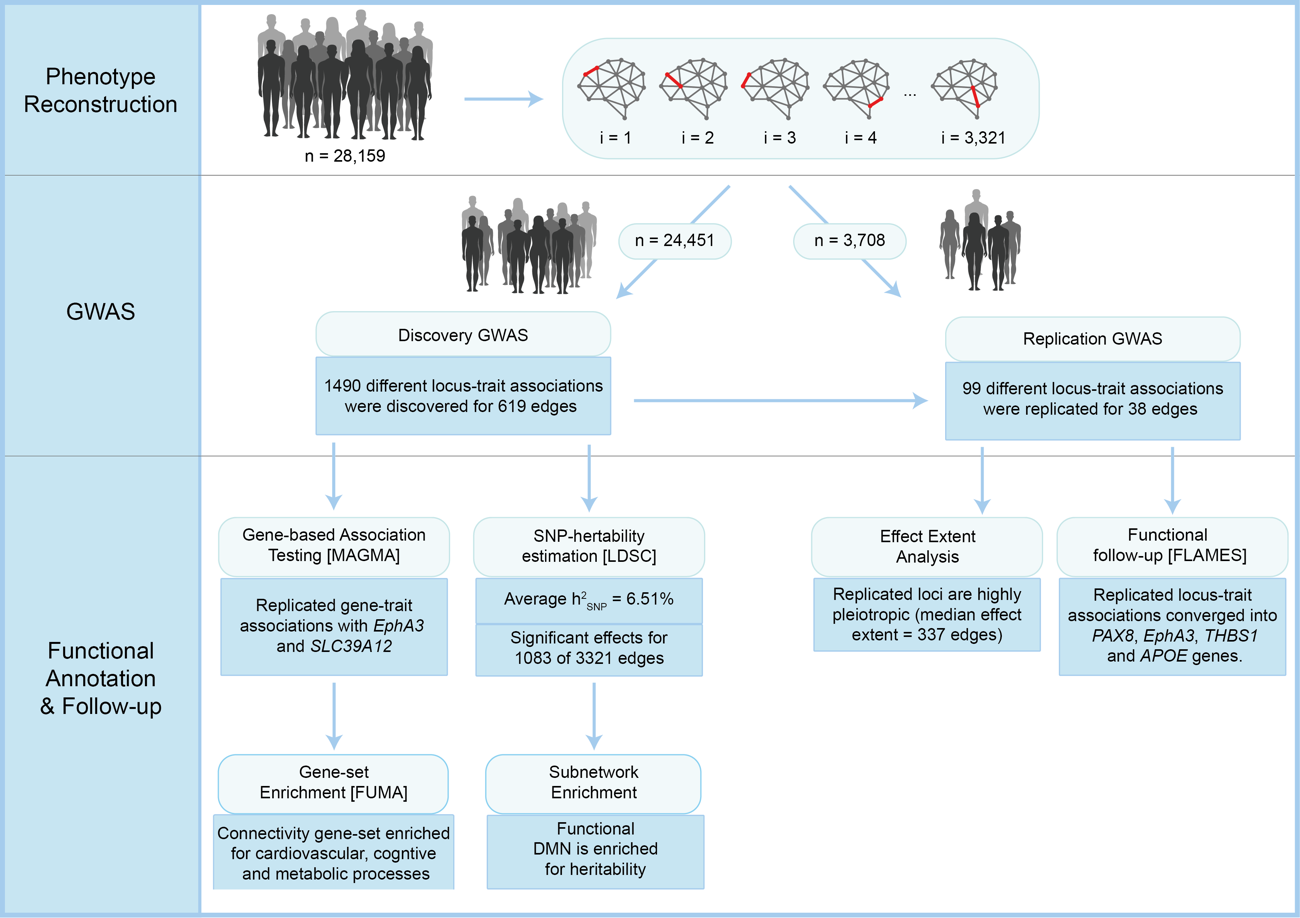

**Supplementary Figure 2. Circos plots of the connections predicted to be associated with the *PAX8* locus**. Only loci significant for an alpha level FDR-corrected for the number of phenotypes tested are drawn.

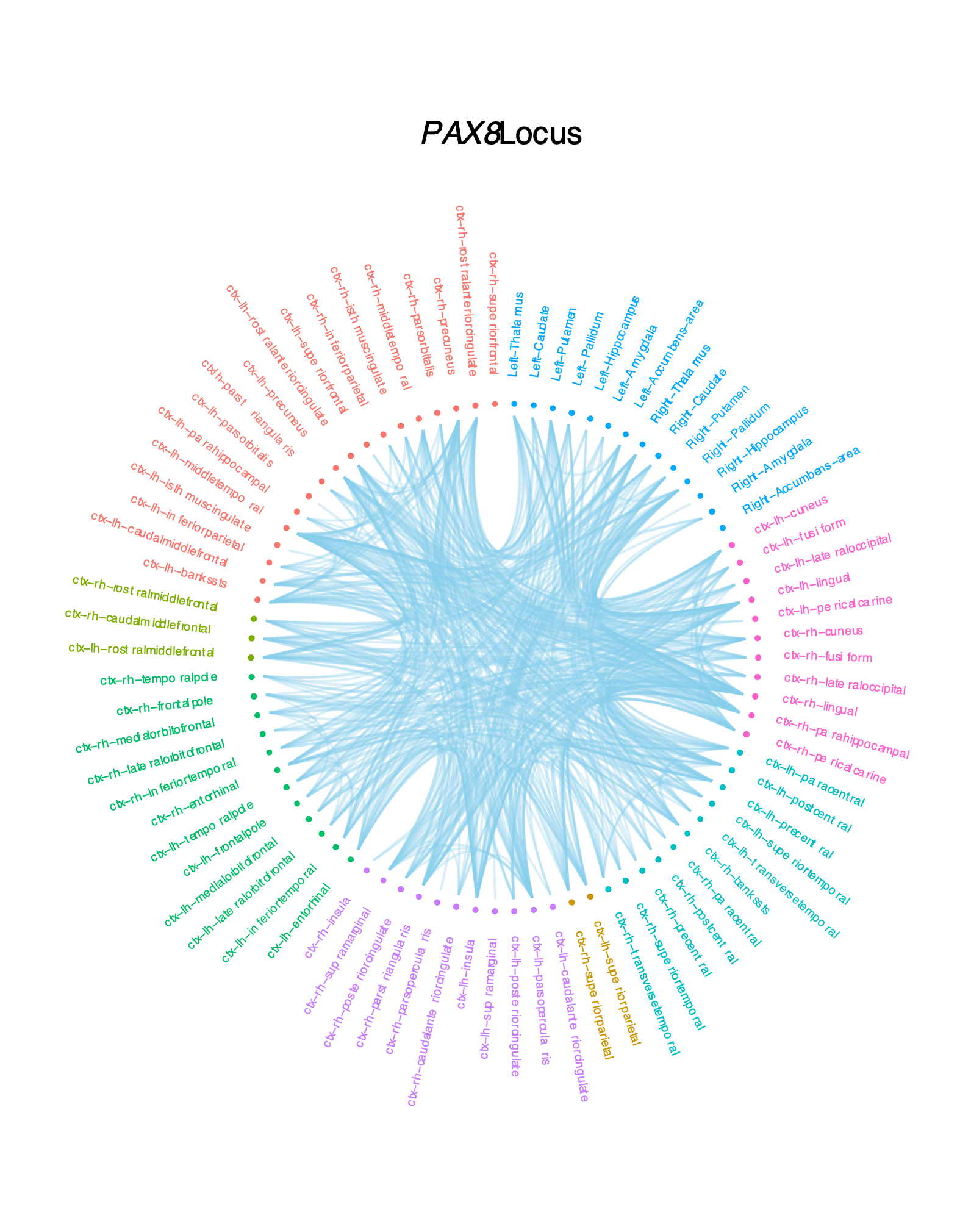

**Supplementary Figure 3. Circos plots of the connections predicted to be associated with the *EphA3* locus**. Only loci significant for an alpha level FDR-corrected for the number of phenotypes tested are drawn.

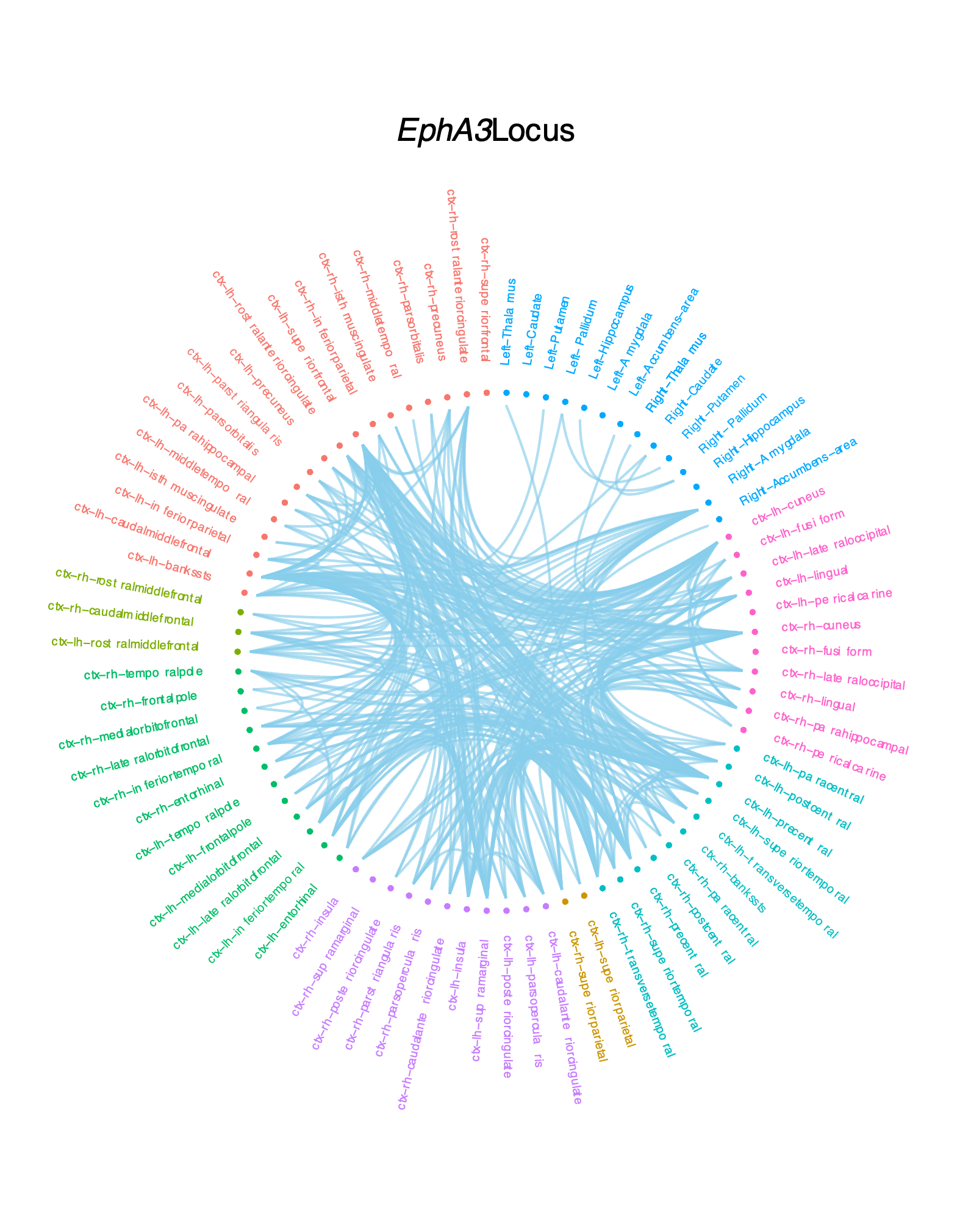

**Supplementary Figure 4. Circos plots of the connections predicted to be associated with the *TBHS1* locus**. Only loci significant for an alpha level FDR-corrected for the number of phenotypes tested are drawn.

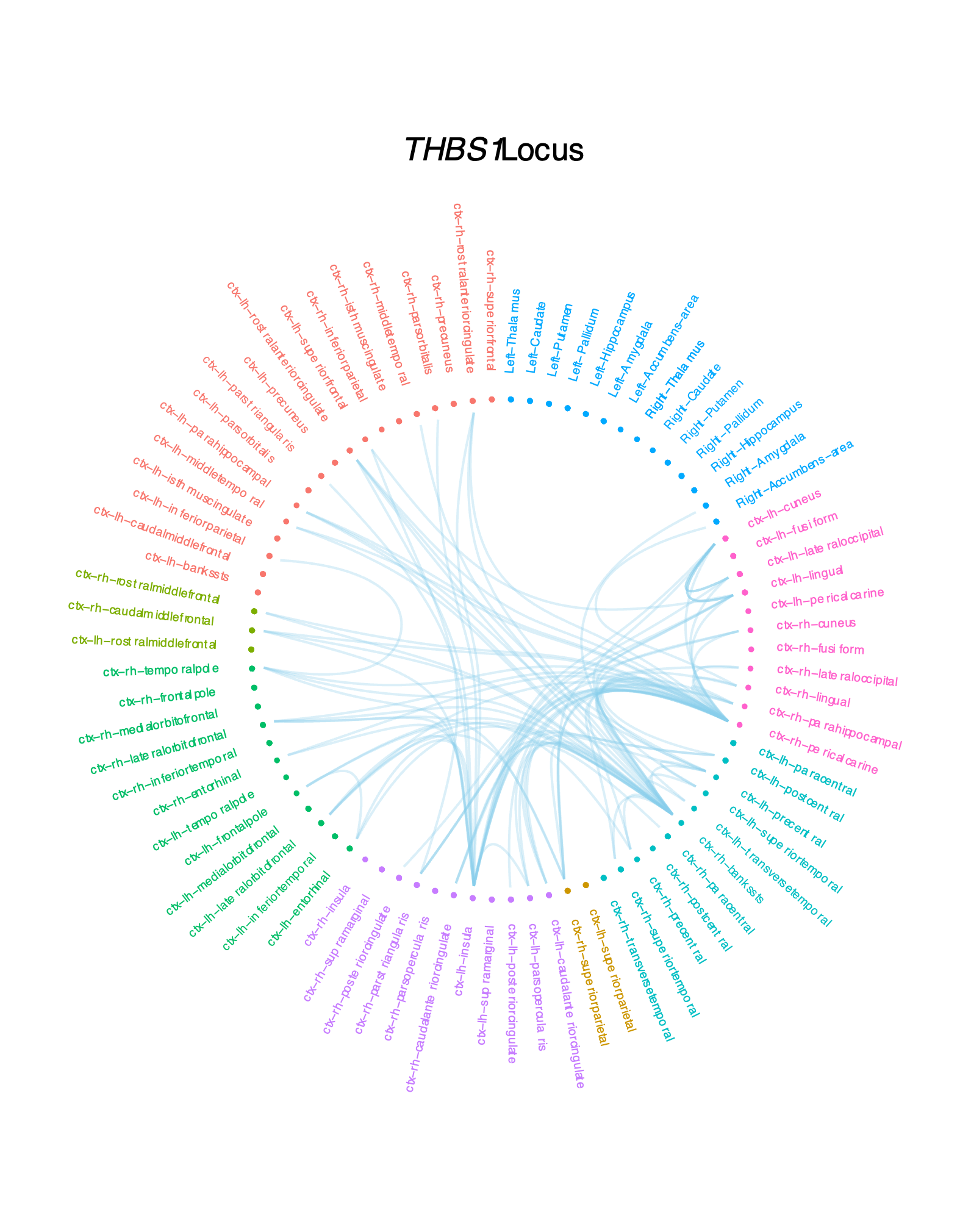

**Supplementary Figure 5. Circos plots of the connections predicted to be associated with the *APOE* locus**. Only loci significant for an alpha level FDR-corrected for the number of phenotypes tested are drawn.

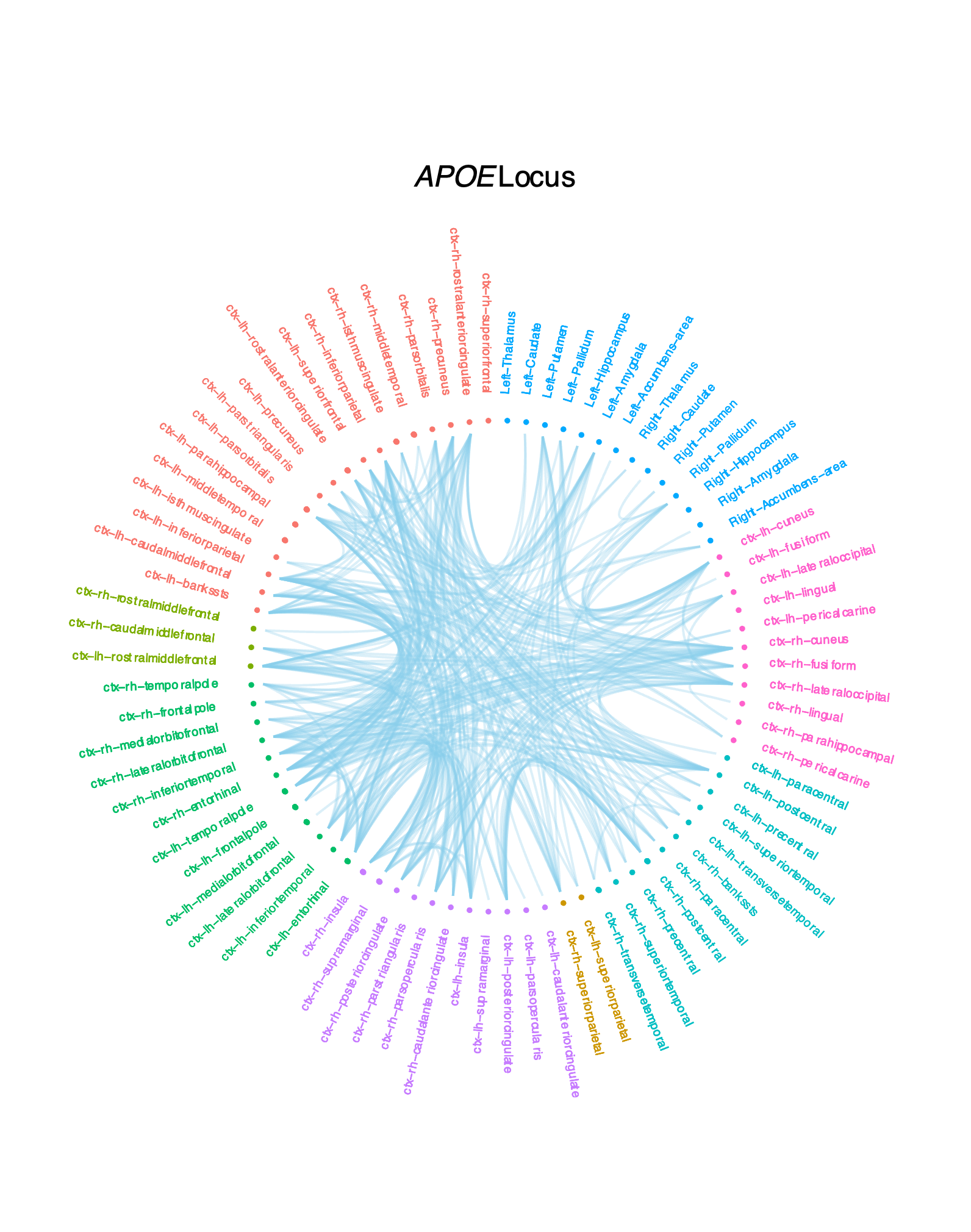

**Supplementary Figure 6. QQ Plots for association of locus across the brain**. Each point represents one different edge. Red line is expectation under no association. Orange line is FDR-correction significance level. The number of points above orange line is the effect extent of the locus throughout the functional connectome. Only loci significant top replicated loci are represented. P-values refer to MAGMA SNP-wise mean locus p-values (**Methods**).

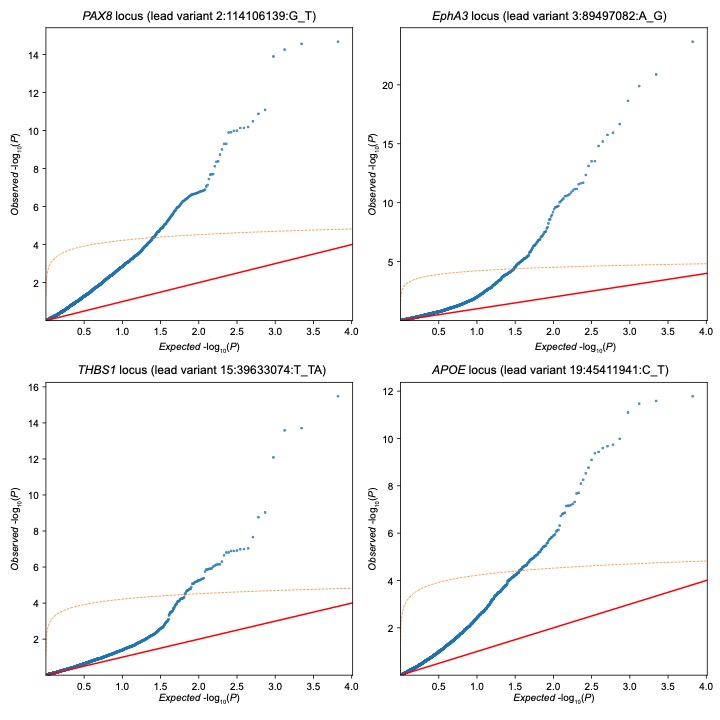

**Supplementary Figure 7. Gene-set enrichment testing for set of replicated genes.**

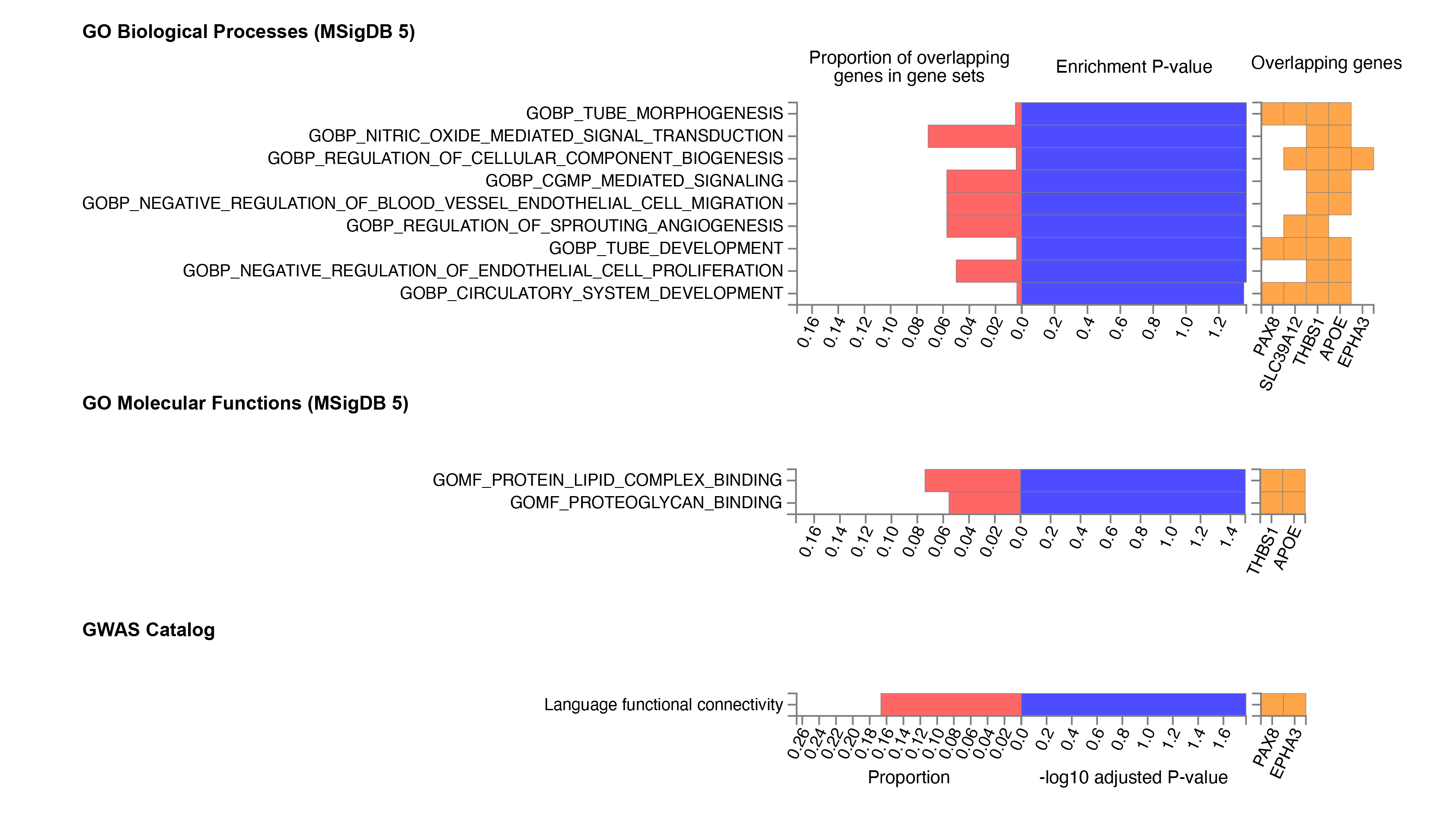

**Supplementary Figure 8. Protein-Protein Interaction Network.** Evidence is derived from experiments, databases, co‑expression, neighborhood, gene fusion and co‑occurrence. Only interacting genes are depicted.

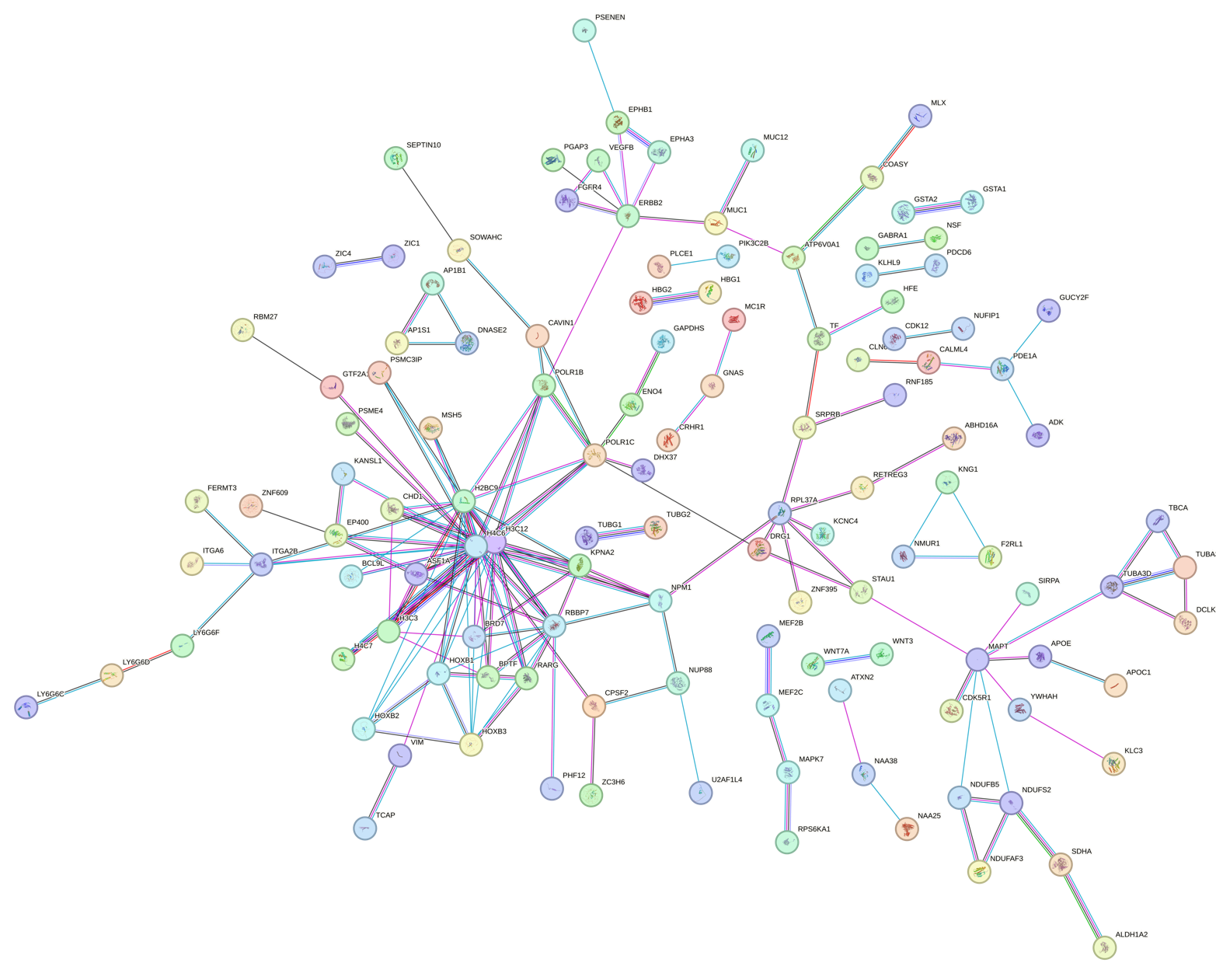

**Supplementary Figure 9. Geneset enrichment testing for complete sex of replicated genes.** Figure continues in the next page.

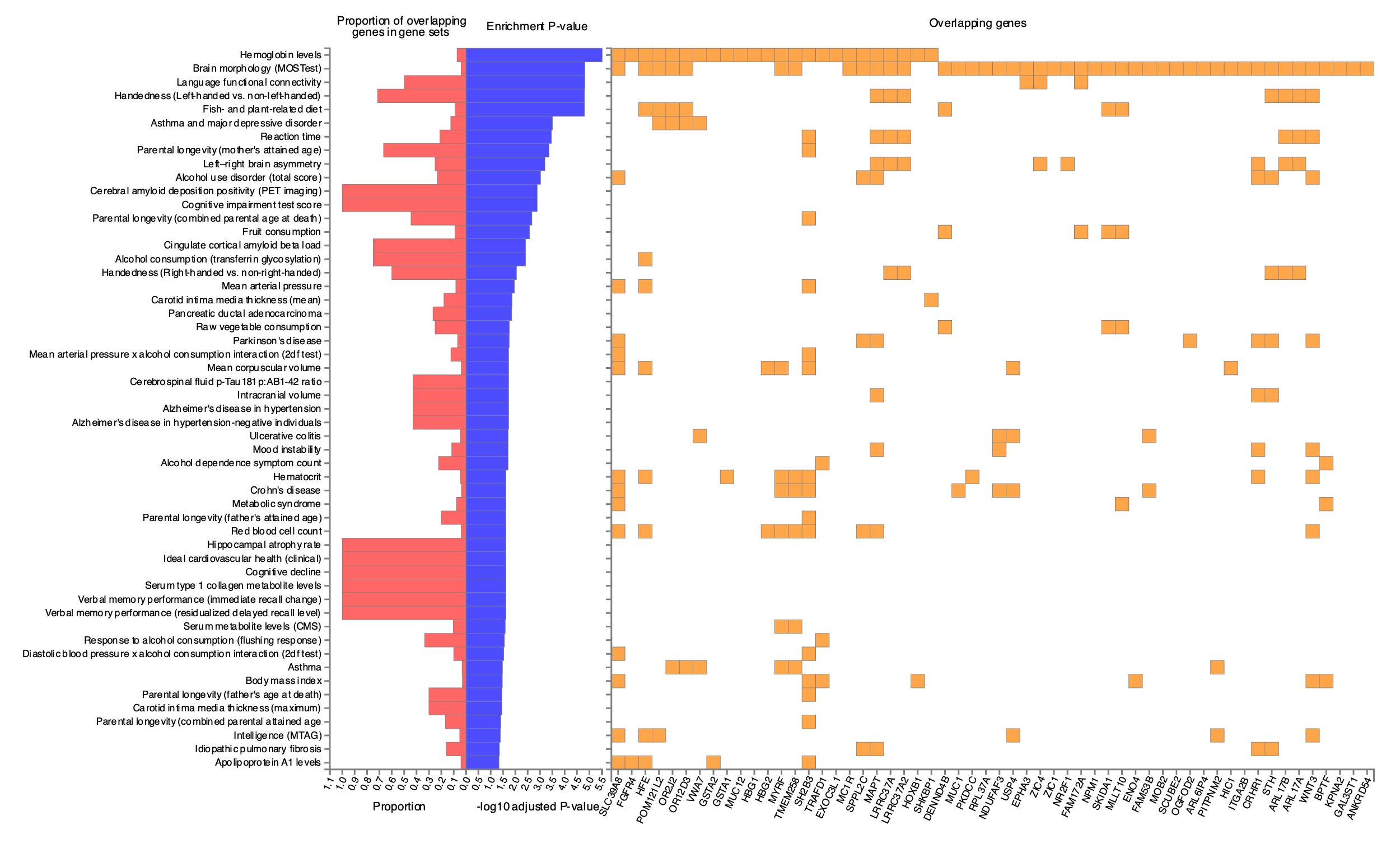

**Supplementary Figure 9. Continued.**

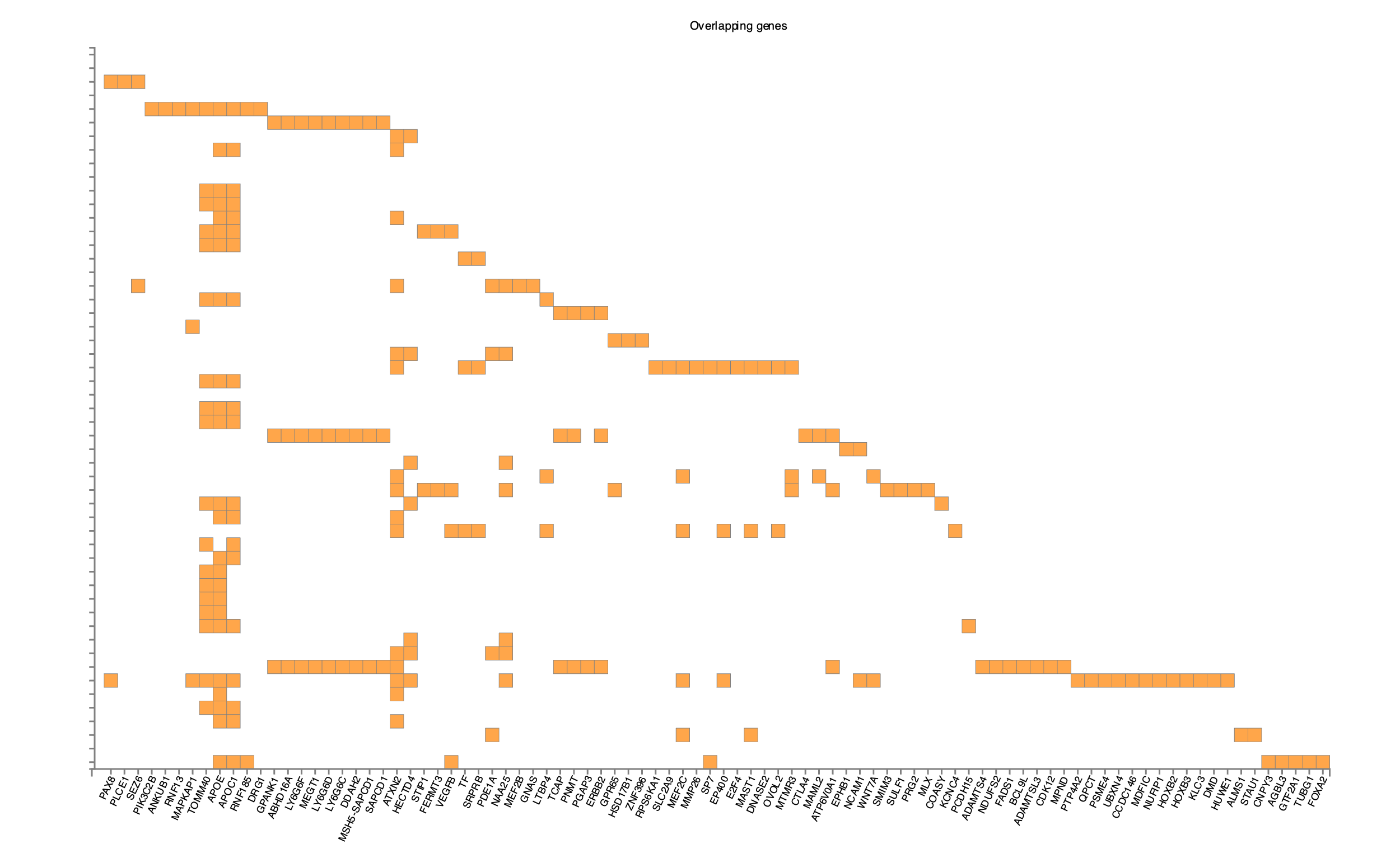

**Supplementary Figure 10. Effect extent of disease genes in the functional connectome.** Each panel represents a gene-set derived from a different case control disease GWAS. Null-models were built per disorder by drawing (i) a random set of brain genes (green), (ii) a LD-aware set of random genes (blue) or (iii) a LD-aware set of brain genes (red); and calculating its mean effect extent. The number of genes drawn equals the number of significant genes for each disorder. Attention Deficit/Hyperactivity Disorder, Anorexia, Autism Spectrum Disorder, Alzheimer’s Disorder, Bipolar Disorder, Depression, Substance Use Disorder, and Schizophrenia. The dashed line is the mean effect extent for the set of disorder genes. P-values are calculated based on the null model with LD-aware set of brain expressed genes.

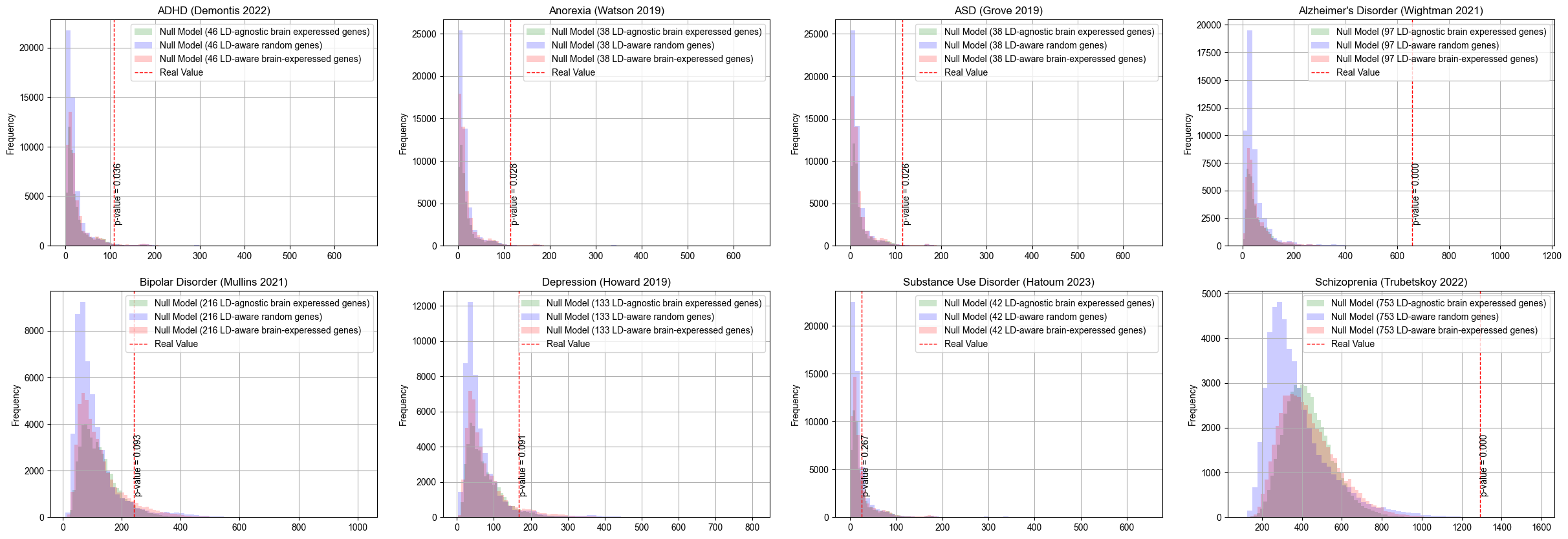

**Supplementary Figure 11. Tissue Expression heatmap of replicated genes**.

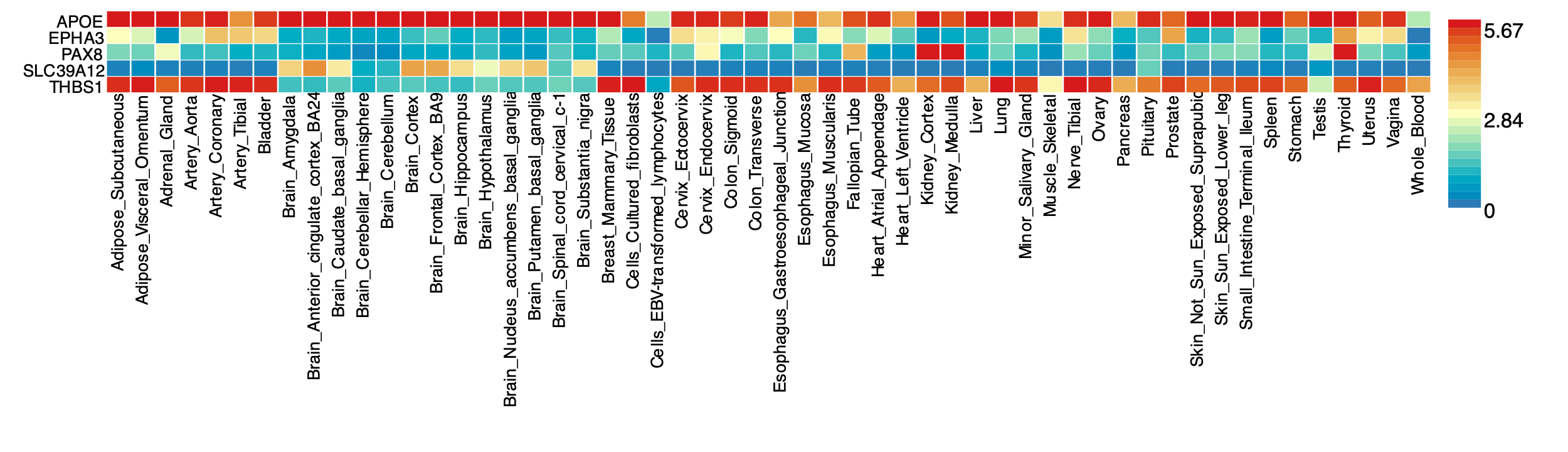

**Supplementary Figure 12. Lifetime expression heatmap of replicated genes.** Pcw: post-conception weeks (prenatal). Yrs: Years (post-natal).

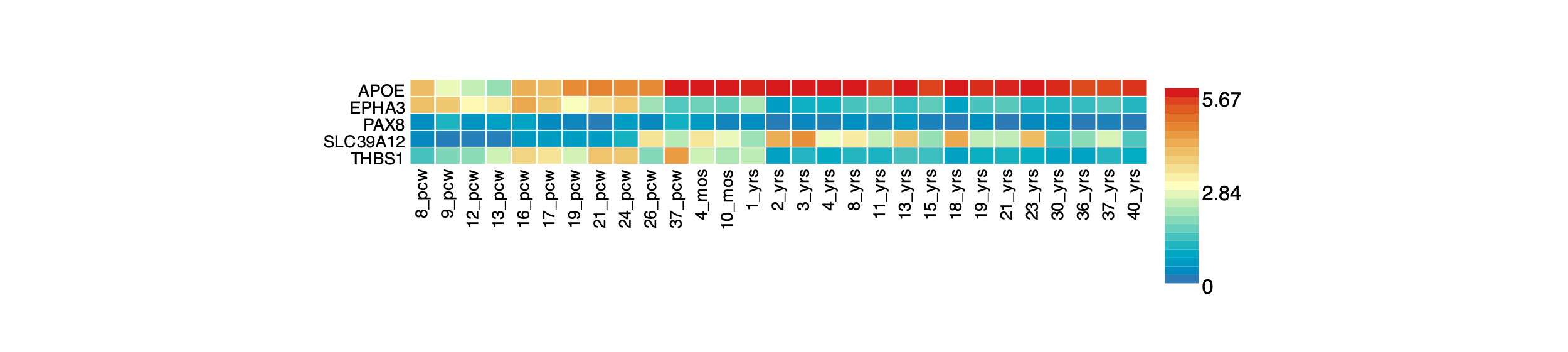
